# Anatomy of aging through organ-resolved multi-modal imaging and deep learning

**DOI:** 10.64898/2026.03.14.26348392

**Authors:** Alec Eames, Dmitrii Glubokov, Alibek Moldakozhayev, Ali Doğa Yücel, Alexander Tyshkovskiy, Kejun Ying, Ludger J. E. Goeminne, Cecília G. de Magalhães, Vadim N. Gladyshev

## Abstract

While aging manifests differently across organs and individuals, existing approaches to measure it lack the spatial resolution to capture this complexity. Here, we develop an approach that applies multi-modal imaging, segmentation algorithms, and deep-learning to assess organ-specific aging across 39 anatomical regions in a total of 134K individuals in the UK Biobank. Our analysis reveals significant organ aging heterogeneity across and within individuals and a remarkable prevalence of organ-specific extreme aging. We validate that our imaging measures capture pathophysiologically meaningful aging through correlation with organ-specific biomarkers, revealing biologically coherent patterns. We find that accelerated organ aging is robustly predictive of corresponding organ disease. We identify the cerebrum as one of the strongest predictors of organismal aging. We investigate organ aging patterns underlying disease risk and find that each disease is linked to aging of highly distinct subsets of organs.

Exploring lifestyle factors and interventions reveals a range of divergent organ-specific effects. Our work establishes a powerful paradigm for noninvasively evaluating human aging at anatomical resolution and population scale.

## Introduction

Aging involves system-wide declines in physiological function and increased vulnerability to mortality and morbidity, stemming from cumulative molecular damage ^1–3^. Yet while aging affects the entire body, distinct patterns of organ disease emerge during aging, highlighting the heterogeneity of the aging process both within and across individuals. Given this variability, assessing aging through comprehensive, organ-specific approaches will be essential for understanding how aging manifests across diverse biological systems, identifying accelerated organ aging before the emergence of pathology, and enabling the development of targeted interventions tailored to specific organs and systems. Together, these applications make high-resolution measures of aging critical for extending healthy human lifespan ^4,5^.

Recent advances in aging research have underscored the value of organ-resolution age measurements, profiling organ aging through physiological parameters ^6^, organ-specific plasma proteins ^5,7^, and image-derived phenotypes (IDPs) ^8^. However, blood-based methods are limited by temporal variability ^9^ and the risk of complications ^10^, while IDP-based approaches rely on a limited number of predefined features that may not fully capture the complex, high-dimensional dynamics of biological aging. Moreover, these methods lack the spatial resolution necessary to comprehensively capture the multifaceted nature of aging.

Medical imaging is an invaluable tool for assessing the physiology and pathology of virtually every major organ in the body ^11,12^. Across modalities, imaging has emerged as a robust marker of biological age, capable of capturing emergent morphological and physiological changes that accompany aging and contribute to disease ^13–16^. Crucially, advances in segmentation algorithms now enable automated extraction of individual organs and tissues from bulk-level images ^17–20^.

Here, we harness multi-modal imaging from the UK Biobank coupled with advanced segmentation methods and deep learning models to noninvasively assess human aging across 39 organs, tissues, and anatomical regions, capturing the aging process at unprecedented spatial resolution for a total of over 130K individuals. Investigating organ-specific aging in the context of disease, mortality, and lifestyle, we uncover a multitude of new insights into the aging process at organ resolution (Figure 1). Our work demonstrates that non-invasive imaging can comprehend organ-specific aging processes at population-level scale, representing a significant step toward achieving widely available personalized aging assessment.

**Figure 1:**
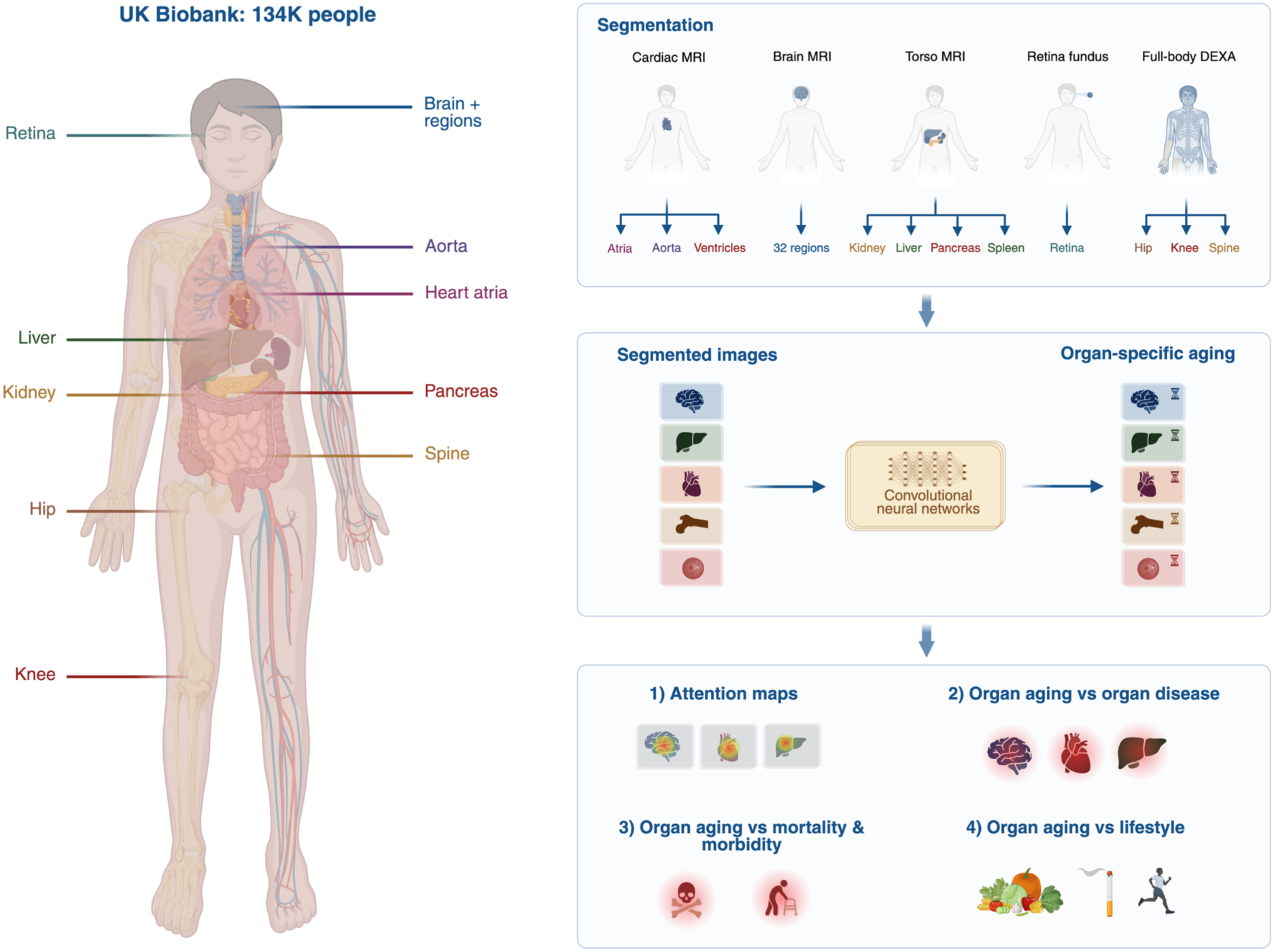
Schematic overview of the study. Beginning with images from a total of 134K people in the UK Biobank, we applied deep learning segmentation methods to isolate specific organs from bulk-level images. These images served as input to convolutional neural networks built to predict organ-specific aging. We assessed the features of organs driving predictions using attention maps, and we explored the relationship between organ aging and disease, mortality, and lifestyle factors.

## Results

### Imaging captures organ-specific aging

Aiming to assess the aging process noninvasively across a wide range of organs, we leveraged five imaging modalities available in the UK Biobank: dual-energy X-ray absorptiometry (DEXA) scans, retina fundus images, and magnetic resonance imaging (MRI) of the torso, brain, and heart. From the DEXA images, we utilized images of the hip, knee, and spine, each representing distinct components of the skeletal system. We also used two full-body DEXA images: one highlighting the skeletal system and another highlighting soft tissue. For the MRI scans, we applied three recently developed segmentation algorithms that were validated, critically, by comparison to gold-standard manual segmentations ^17–21^. This enabled accurate segmentation of various organs and regions from bulk-level images. Specifically, we segmented 32 brain regions from the brain MRIs; the liver, kidneys, spleen, and pancreas from the torso MRIs; and the heart ventricles, atria, and aorta from the cardiac MRIs (Figure 2A). The number of available images varied by modality, ranging from ∼29K for brain MRI regions to ∼90K for retina fundus images, with DEXA and torso MRI having intermediate numbers of images (∼75-80K) (Table S2A).

**Figure 2:**
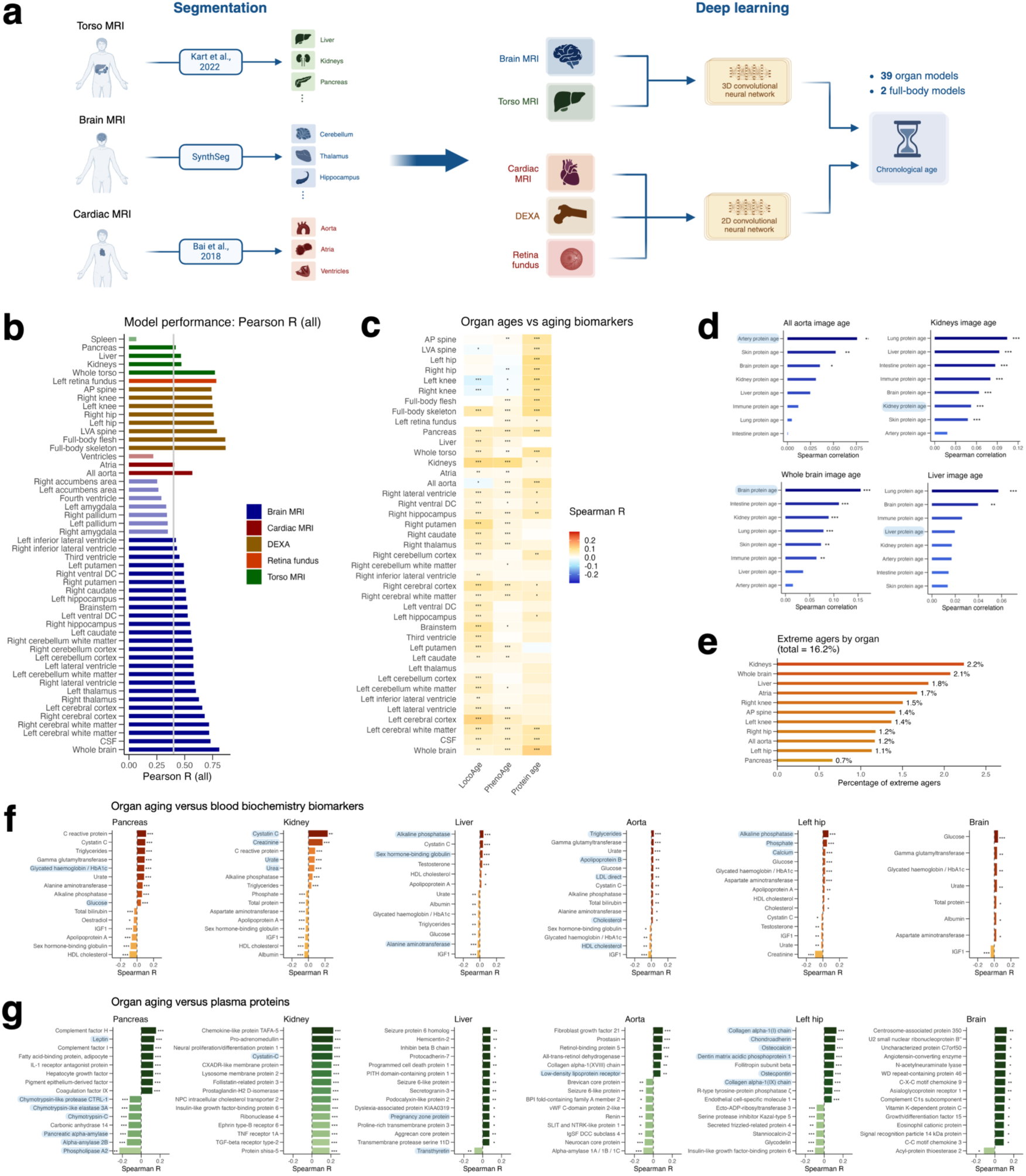
Organ imaging models overview. **(a)** Schematic depicting segmentation process for torso, brain, and cardiac MRIs, and overview of model training for each image type. **(b)** Correlation performance (Pearson R) of each model in predicting age across all out-of-fold participants. Retained models exceed the 0.4 Pearson R threshold. **(c)** Association between age deviation of image-based organ age and PhenoAge, LocoAge, and plasma protein-based age deviation. **(d)** Comparison between protein-based and image-based organ age deviation for the 4 organs that correspond between modalities: artery / aorta, kidneys, brain, and liver. **(e)** For a subset of 11 non-overlapping organs, bar plot depicting the proportion of people exhibiting organ-specific extreme aging. **(f)** For a subset of major representative organs, correlation between organ age deviation and blood biochemistry parameters (top 15 that were statistically significant shown here). **(g)** For a subset of major representative organs, correlation between organ age deviation and plasma protein levels (top 15 proteins that were statistically significant shown here). Adjusted p-value < 0.001 = ***, adjusted p-value < 0.01 = **, adjusted p-value < 0.05 = *.

We then developed two convolutional neural networks (CNNs) to predict age: a 2D CNN designed for 2D images, including retina, DEXA, and cardiac MRI images, and a 3D CNN designed for the 3D torso and brain MRI images (Figure 2A). For each image type, we trained a model to predict chronological age, training with participants who were generally healthy at baseline (see Methods). Metadata features of the training and full imaging cohort are plotted in Figure S1D and S1F. For both cohorts, ages range from ∼40-85 years. We evaluated the accuracy of our models using cross-validation and retained models with a cross-validated Pearson correlation (R) between predicted and actual age of at least 0.4, yielding 41 final models: 39 specific to organs and regions––including the whole-brain and whole-torso images––and 2 full-body models (Figure 2B). The accuracy of these models in predicting chronological age, as measured by Pearson correlation, ranged from 0.41 to 0.87 (Figure 2B). Models that did not meet this performance threshold were primarily from small anatomical structures with limited image resolution. We then compared age estimates across organs by computing the correlation between the age deviation (predicted minus chronological age) of each pair of organs. Overall, we observed statistically significant correlations, with stronger associations between related organs (Figure S1B).

To validate our newly developed image aging models against established gold-standard biological aging metrics, we compared our organ age deviation estimates to several well-validated quantitative indicators of the biological aging process, or aging biomarkers, ranging from clinical and functional to molecular (Figure 2C-D, Figure S1E). These included PhenoAge, a widely used aging biomarker based on clinical parameters strongly linked to mortality and morbidity ^22^, LocoAge, which captures age-related functional decline through locomotor activity patterns ^23^, and a plasma protein-based aging signature developed by Goeminne et al., 2025 that reflects systemic molecular aging processes ^7^. For each biomarker, we computed the correlation between the predicted age deviation (difference from chronological age) and the age deviation of our organ image models and found statistically robust correlations across the majority of organ image models.

We next compared our organ age deviations to plasma-protein-based organ age estimates ^7^, which revealed striking organ-specific associations: aorta image age correlated most strongly with artery protein age, brain image age with brain protein age, and, albeit weaker, kidney image age with kidney protein age; however, liver aging did not associate across modalities (Figure 2D, Figure S1E).

To further validate our models, we compared our organ age deviations to polygenic risk scores for organ-specific age-related disease (Figure S3D), which quantify an individual’s disease risk based on genetic variants observed in their genome ^24^. This analysis revealed remarkable organ-specific associations: Alzheimer’s risk scores correlated most strongly with brain aging, ischemic stroke risk with aorta and cardiac aging, osteoporosis risk with skeletal aging, and type 2 diabetes risk with pancreas aging. Similarly, macular degeneration risk scores showed strong associations with retinal aging, and Parkinson’s disease correlated most with aging of the brain’s left inferior lateral ventricle. We also observed some unexpected patterns, including negative correlations between Parkinson’s risk scores and certain brain regions’ aging, and cardiovascular disease risk scores associating most strongly with spinal aging.

We then examined the prevalence of individuals displaying extreme aging in a single organ. We defined extreme organ aging as an age deviation exceeding two standard deviations above the mean in only one organ for a subset of 11 non-overlapping organs within the same individual. Remarkably, we found that nearly 1 in 6 participants displayed extreme organ-specific aging (16.2%), which was most commonly observed in the kidney and brain (Figure 2E). The age distribution of extreme agers compared to non-extreme agers was similar (Figure S1G).

To gain insight into the biological features underlying image-based organ aging, we correlated organ aging estimates with blood biochemistry parameters, plasma proteins and metabolites, and physiological features (Figure 2F-G, Figure S2, Table S3). Across organs, this revealed a range of noteworthy associations.

Pancreas aging correlated with markers of glycemic control (glucose, HbA1c) and pancreatic exocrine function (digestive enzymes like pancreatic alpha-amylase), kidney aging with well-validated markers of renal function (cystatin C, creatinine) ^25^, and liver aging with liver function markers including alkaline phosphatase and liver-derived proteins such as sex hormone-binding globulin and transthyretin ^26,27^. Aorta aging was linked most strongly to atherogenic lipids and lipoproteins (triglycerides, Apolipoprotein B, LDL cholesterol, and VLDL-derived lipids) ^28^ as well as functional measures of cardiovascular health such as blood pressure and heart rate, while skeletal aging correlated with both bone-related minerals (phosphate, calcium) as well as skeletal system-derived proteins (collagen proteins, chondroadherin, osteocalcin, alkaline phosphatase). Brain aging corresponded to markers of metabolic health, inflammatory cytokines, and proteins such as GDF15, whose levels have been causally linked to Alzheimer’s disease development ^29^ (Table S3).

To further investigate the molecular mechanisms underlying organ aging, we performed gene set enrichment analysis for a range of organs using plasma protein levels and MSigDB hallmark pathways ^30,31^ (Figure S3C), revealing both universal and organ-specific aging signatures that align with established hallmarks of aging. The most consistent signal, observed across nearly every organ, was enrichment of inflammatory pathways (e.g., complement, interferon, and IL6/JAK/STAT signaling), a well-established hallmark of aging ^32^, highlighting age-related chronic inflammation as a unifying mechanistic driver of organ aging. Conversely, some pathways displayed selective organ-specific enrichment––mTORC1 signaling was enriched only in cardiovascular tissues and the spine, while TGF-beta, a major driver of age-related fibrosis ^33,34^, was significantly enriched only in the heart––providing insights into distinct aging processes across different organ systems.

### Attention maps highlight spatial features driving age predictions

We sought to gain insights into the features driving our models’ predictions by constructing attention maps with the Grad-CAM framework ^35^. While the small resolution of many images limited intra-organ analysis, our models generally highlighted biologically meaningful regions (Figure 3, Figure S4).

**Figure 3:**
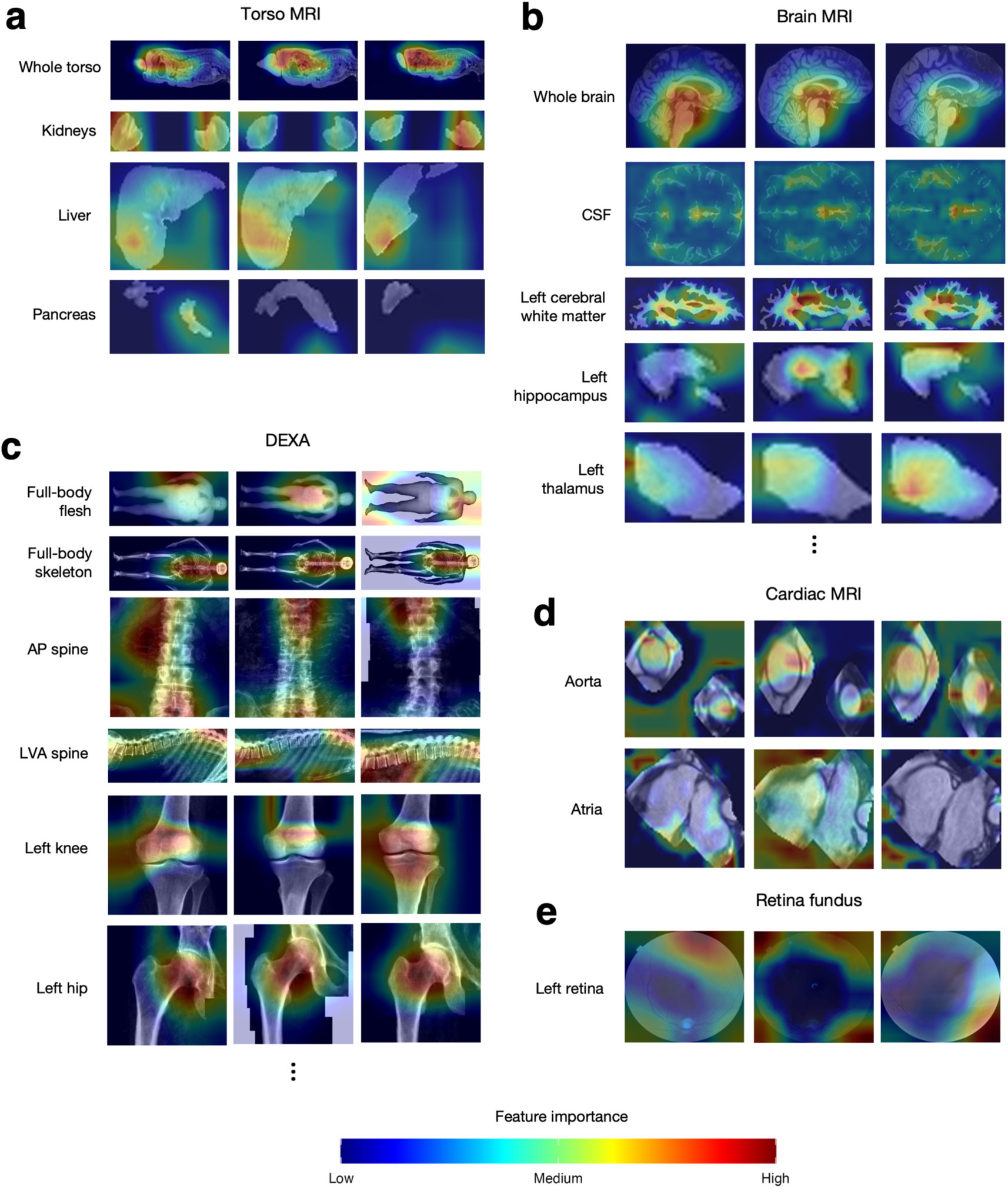
Attention maps overview. Attention maps displaying representative images of three individuals (columns) for the torso MRI models **(a)**, selected brain MRI models **(b)**, selected DEXA models **(c)**, cardiac MRI models **(d)**, and retina fundus models **(e)**. For the 3D images (torso and brain MRI), all images are taken from the middle slice of the axial dimension except for the whole-torso and whole-brain images, which are taken from the middle slice of the sagittal dimension.

Analysis of the larger-resolution images revealed several noteworthy features. The whole-torso aging model highlighted the thorax (Figure 3A), a region containing critical cardiorespiratory organs––the heart and lungs––whose dysfunction accounts for a large fraction of age-related disease and death ^36^. In the brain, our model particularly emphasized aging patterns in the subcortical regions (Figure 3B), suggesting these areas undergo more prominent morphological shifts with age than cortical regions.

The skeletal models revealed several notable patterns. Consistent with the torso MRI model, the two full-body DEXA models highlighted the upper body, including the thorax, as being most salient for age prediction. The full-body skeleton model also highlighted the skull particularly strongly (Figure 3C), and the full-body skeleton model’s attention was most concentrated on the spine. For both the hip and knee models, attention maps highlighted the joints as being most critical for predicting aging, suggesting the joints––more so than the bones themselves––may be particularly vulnerable to structural alterations as aging takes place. We describe the full interpretation of each attention map in Table S4.

### Organ aging predicts corresponding organ disease

We examined the relationship between organ aging and the future risk of developing 25 major age-related diseases (Figure 4A), excluding individuals already diagnosed with the disease or diagnosed within a year of imaging. Our analysis revealed striking associations between aging of organs and diseases corresponding to those organs across a wide range of cases (Figure 4B-F).

**Figure 4:**
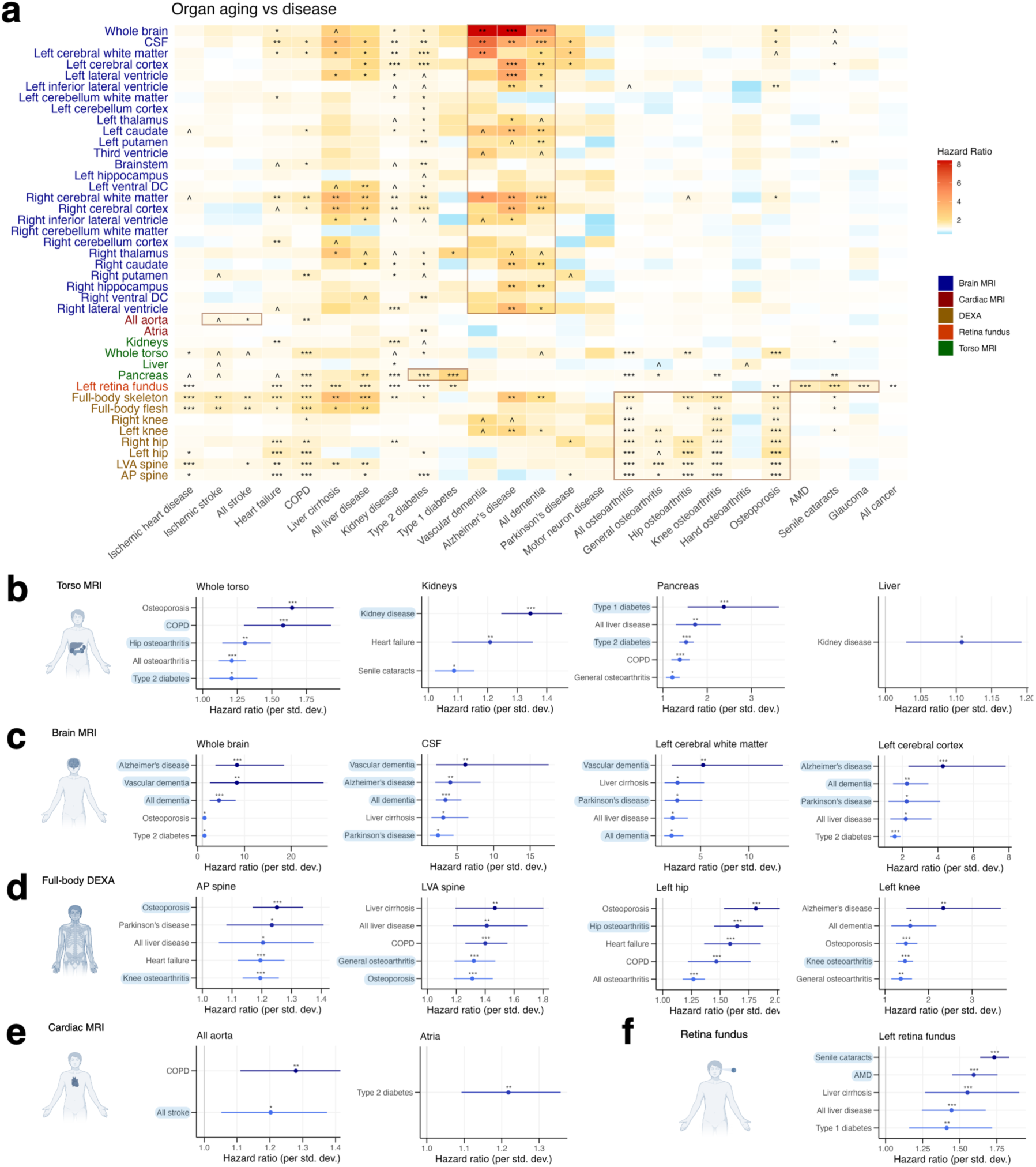
Association between organ aging and major disease. **(a)** Heatmap depicting the hazard ratio and significance between each organ and each major disease, adjusting for chronological age and sex (adjusted p-value < 0.001 = ***, adjusted p-value < 0.01 = **, adjusted p-value < 0.05 = *, adjusted p-value < 0.1 = ^). Forest plots depict the top 5 most strongly associated diseases that were statistically significant, ranked by hazard ratios, for torso MRI models **(b)**, brain MRI models **(c)**, DEXA models **(d)**, cardiac MRI models **(e)**, and retina fundus models **(f)**. Diseases related to a particular organ or region are highlighted.

For example, torso aging was most strongly associated with torso diseases, including chronic obstructive pulmonary disease (COPD), hip arthritis, and type 2 diabetes. Kidney aging was most strongly linked to kidney disease, and pancreas aging to type 1 diabetes, followed closely by type 2 diabetes (Figure 4B). Similarly, brain aging displayed robust and specific associations with Alzheimer’s disease and vascular dementia (Figure 4C).

In the skeletal system, skeletal aging was associated with both osteoporosis and osteoarthritis. We also observed remarkable region-specific associations: hip aging was strongly predictive of hip arthritis, and knee aging of knee arthritis (Figure 4D). Aortic aging displayed a strong association with stroke risk (Figure 4E), and retinal aging was most predictive of ocular conditions—cataracts, age-related macular degeneration (AMD), and glaucoma (Figure 4F, Figure 4A).

Beyond organ-specific patterns, we observed several further noteworthy results. For instance, eye aging was associated with a range of non-ocular diseases, including ischemic heart disease, COPD, and kidney disease, suggesting the eye may serve as a biomarker of aging and disease throughout the body (Figure 4A). We also observed notable inter-organ relationships: kidney aging was linked to heart failure, and pancreas aging to kidney disease. Further, pancreas aging was strongly associated with liver disease (Figure 4B). While some of these cross-organ associations may reflect pathophysiological connections, others likely represent correlated age-related dysfunction across multiple organ systems, and we caution against inferring causality from these associations.

Two models produced unexpected results. Atrial aging was predictive only of type 2 diabetes (Figure 4E), and liver aging predicted only kidney disease (Figure 4B). While the particularly low resolution of the atria images likely impeded the model’s ability to capture meaningful features, the absence of liver aging’s association with most diseases is consistent with its conspicuous aging resilience. In contrast to other abdominal organs, the liver displays remarkably minimal functional decline as it ages ^37^. Our model, therefore, may capture age-related morphological changes that are benign structural alterations rather than indicators of dysfunction, which may explain its minimal association with liver disease and other diseases.

### Cerebrum aging strongly associates with mortality and morbidity

We sought to identify which organs’ aging was most relevant to organismal aging, as reflected by all-cause mortality and age-related morbidity (see Methods). Notably, we found that the brain––particularly the cerebral cortex, cerebral white matter, and cerebrospinal fluid (CSF) patterns––was among the organs whose aging was most strongly linked to all-cause mortality risk (Figure 5A). Similarly, cerebral aging showed a robust association with morbidity risk, although here, it was comparable to the full-body aging models (Figure 5B). Pancreas aging demonstrated stronger associations with mortality and morbidity risk than kidney aging, which, in turn, showed stronger associations than liver aging. Similarly, hip aging was much more closely linked to morbidity than knee aging. Remarkably, aging of the cerebellum––across both the cortex and white matter––displayed no association with either mortality or morbidity (Figure 5A-B).

**Figure 5:**
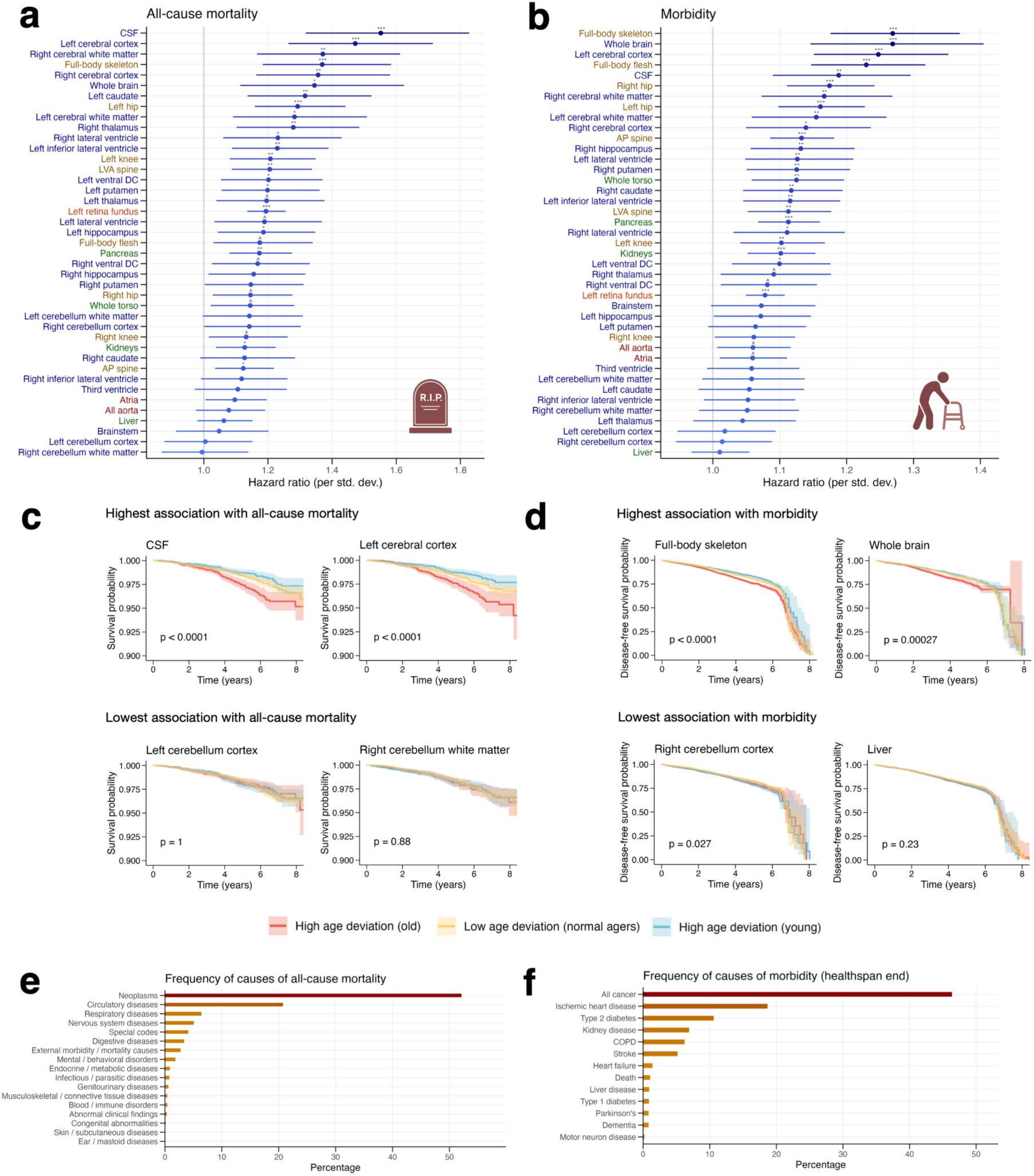
Association between organ aging and all-cause-mortality and morbidity. **(a)** Forest plots depicting each organ’s association with all-cause mortality, controlling for chronological age and sex. **(b)** Forest plots displaying each organ’s association with morbidity, controlling for chronological age and sex. **(c)** Kaplan-Meier curves depicting survival probability for individuals with high age deviation (old) (> 75th percentile), low age deviation (normal agers) (25th to 75th percentile), and high age deviation (young) (< 25th percentile) for the two organs with the highest association with all-cause mortality and the two organs with the lowest association with all-cause mortality. **(d)** Kaplan-Meier curves depicting major-disease-free survival probability for the two organs with the highest association with morbidity and the two organs with the lowest association with morbidity, stratified by the same groups as in (c). **(e)** For subjects with imaging data, bar plot depicting the frequency of causes of all-cause mortality. **(f)** For subjects with imaging data, bar plot depicting the frequency of causes of morbidity. Adjusted p-value < 0.001 = ***, adjusted p-value < 0.01 = **, adjusted p-value < 0.05 = *, adjusted p-value < 0.1 = ^.

To validate these findings, we generated survival curves stratified by baseline age deviation for organs with the strongest and weakest associations with mortality and morbidity. These curves highlighted our previous observations: baseline age deviation in the CSF and cerebral cortex was linked to starkly divergent survival trajectories in the following years, and age deviations in the total body skeleton and brain models strongly predicted the risk of major disease development in subsequent years. In contrast, baseline age deviation in the cerebellum did not lead to marked differences in survival or morbidity trajectories (Figure 5C-D).

Making these findings especially noteworthy is the fact that brain diseases constitute only a small fraction of major mortality and morbidity causes and are dwarfed in prevalence by conditions like cancer and ischemic heart disease (Figure 5E-F). Despite this, aging of the brain is among the most powerful predictor of these organismal-wide aging outcomes.

### Age-related diseases follow from aging of distinct sets of organs

Aging is the leading risk factor for chronic disease, yet how organ aging leads to organ-specific disease remains incompletely understood. While the analysis in Figure 4 examined which diseases each organ aging measure predicted most strongly, we next addressed the complementary question: for specific diseases, which organ aging measures are the most predictive? To investigate this relationship, we analyzed which organs’ aging patterns were most predictive of disease onset across several major conditions that affected organs captured in our models.

Our analysis revealed both organ-specific and multi-organ patterns of disease prediction. For Alzheimer’s disease, we observed associations with aging for only a subset of brain regions. The cerebrum, lateral ventricles, and CSF showed the strongest associations with Alzheimer’s risk, while the caudate, hippocampus, and thalamus displayed weaker links. Notably, similar to mortality and morbidity risk, cerebellum aging did not predict Alzheimer’s disease onset. Aging of the brainstem, putamen, and ventral diencephalon was also not associated with Alzheimer’s disease (Figure 6A).

**Figure 6:**
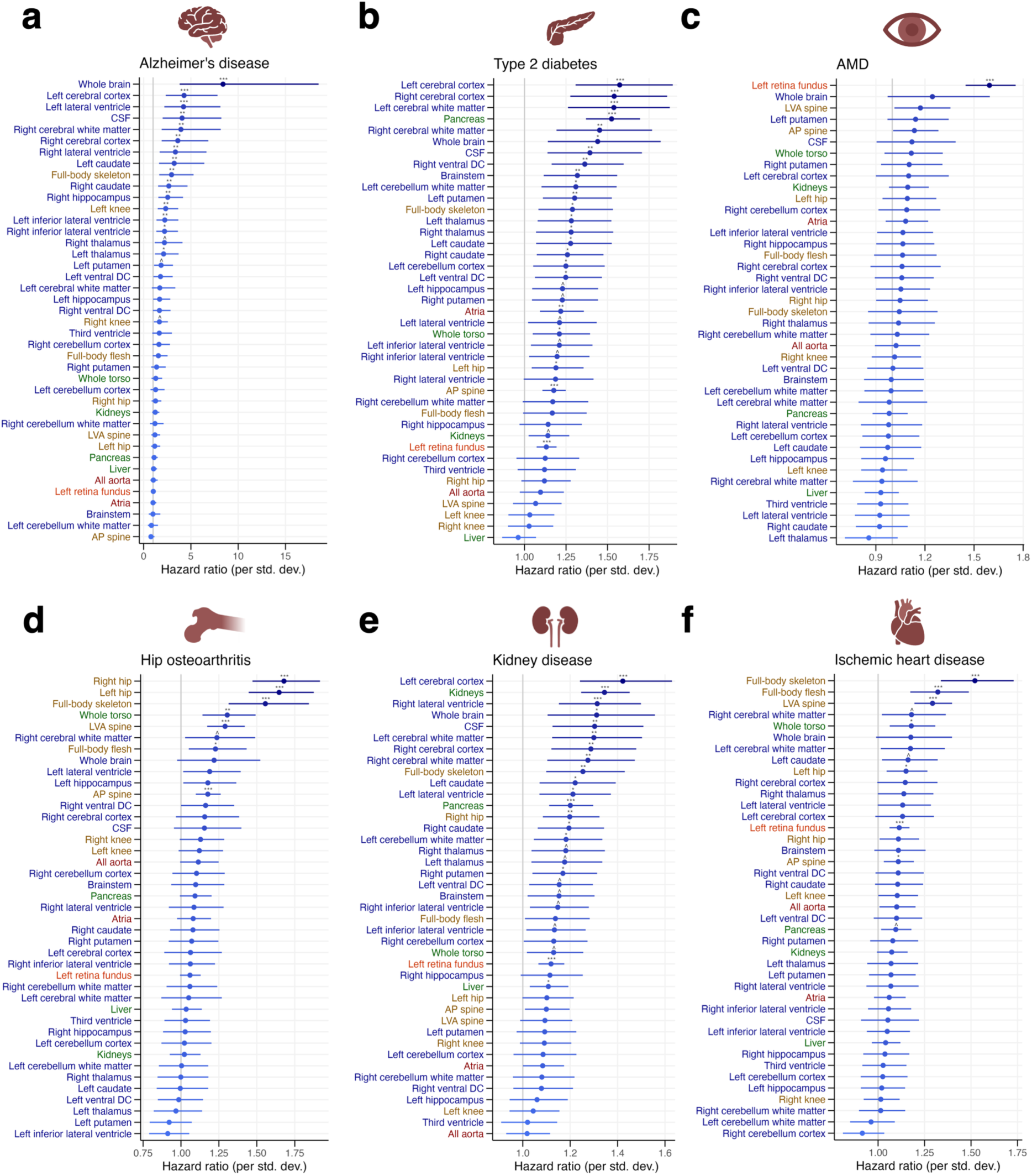
**Relationship between age-related disease and organ aging**. Forest plots depict the hazard ratios between organ aging and disease, adjusting for chronological age and sex for the following diseases: Alzheimer’s disease **(a)**, type 2 diabetes **(b)**, age-related macular degeneration (AMD) **(c)**, hip osteoarthritis **(d)**, kidney disease **(e)**, and ischemic heart disease **(f)**. Adjusted p-value < 0.001 = ***, adjusted p-value < 0.01 = **, adjusted p-value < 0.05 = *, adjusted p-value < 0.1 = ^.

Type 2 diabetes demonstrated a distinct pattern. While pancreas aging was among the top predictors of risk––in alignment with the pancreatic beta cell dysfunction characteristic of the disease ^38^––aging across multiple organs also showed significant associations. Soft tissue aging was generally more predictive than skeletal components like the hip and knee, and brain aging showed a particularly strong association (Figure 6B). Similarly, kidney disease was strongly predicted by kidney aging but was also associated with aging of a range of organs, particularly the brain. (Figure 6E).

In contrast, some conditions showed highly specific organ-aging associations. Age-related macular degeneration (AMD) was significantly linked only to aging of the retinal fundus, suggesting AMD emerges solely from aging of the retina (Figure 6C). Similarly, hip arthritis was best predicted by hip aging, with aging of other organs displaying substantially lower hazard ratios (Figure 6D).

In alignment with the systemic nature of the disease, ischemic heart disease was best predicted by the full-body models (the full-body skeleton and full-body flesh models were the first and third best predictors, respectively) (Figure 6F). Notably, heart disease was better predicted by the full-body skeleton model than the full-body flesh model. This finding may be explained by skeletal aging’s stronger associations with cardiovascular risk markers including C-reactive protein, HbA1c, and triglycerides ^28,39^ (Table S6) as well as central adiposity measures such as waist-to-hip ratio (Table S7, Figure 7E), suggesting that our skeletal aging model captures systemic metabolic and inflammatory processes more directly linked to heart disease risk than soft-tissue aging.

**Figure 7:**
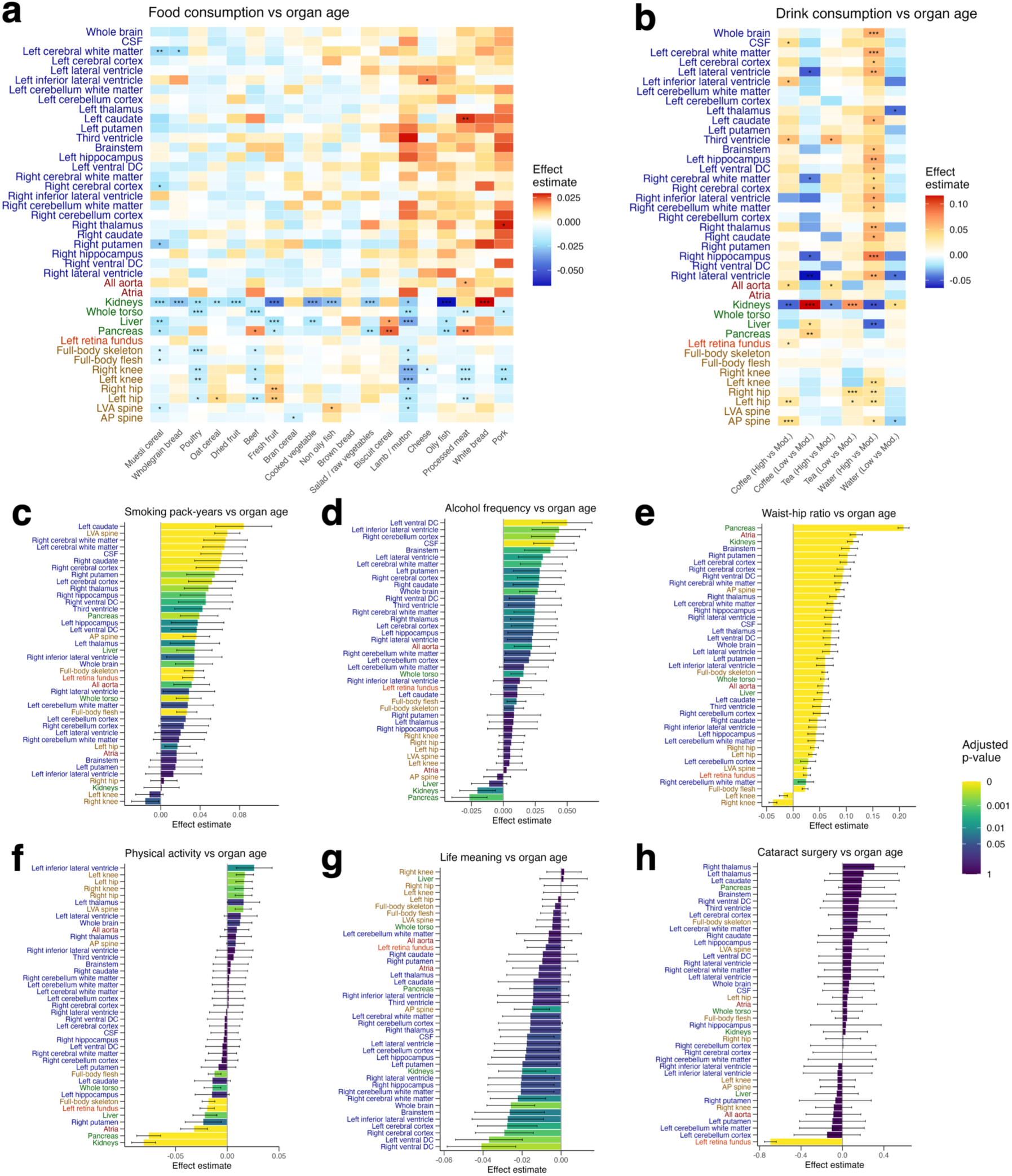
Association between organ aging and lifestyle factors and interventions. **(a)** Association between organ aging and consumption of different foods (adjusted p-value < 0.001 = ***, adjusted p-value < 0.01 = **, adjusted p-value < 0.05 = *). **(b)** Association between organ aging and intake of coffee, tea, and water. Association between organ aging and smoking **(c)**, alcohol frequency **(d)**, waist / hip ratio **(e)**, physical activity level (IPAQ score) **(f)**, life meaning **(g)**, and cataract surgery **(h)**.

To gain insights into how inter-organ communication and coordination may contribute to disease during aging, we investigated whether anatomically remote organ aging measures provided additional predictive value beyond local organ-specific aging for disease prediction. Across six major diseases, we found that multiple remote organs provided statistically significant improvements in predictive performance when added to local organ models (Figure S5). Consistent with the results in Figure 6, we found that aging of several brain regions, particularly the cerebrum, improved predictive performance for type 2 diabetes and kidney disease (Figure S5B, S5E). Hip and knee arthritis were predicted better by the addition of aging metrics for other skeletal regions, such as the spine (Figure S5C-D), and Alzheimer’s and heart disease similarly benefited from incorporating skeletal aging measures (Figure S5A, S5F).

Together, these patterns suggest that certain organ diseases may emerge as a consequence of coordinated age-related processes across anatomically distinct systems.

### Diverse lifestyle factors and interventions are linked to organ-specific aging patterns

While lifestyle factors are known to influence aging trajectories and risk of health outcomes ^40–42^, how they relate to aging at the level of individual organs remains unclear. To explore this, we investigated the association between organ aging and a wide range of factors––including diet, smoking, alcohol, and obesity––as well as interventions such as cataract surgery. Given the observational nature of these analyses, we emphasize that no causal conclusions should be drawn from these results.

Among dietary factors, we found that meat consumption was associated with reduced aging in the skeletal system, particularly the hips and knees, suggesting a potential link between dietary protein intake and skeletal aging. Additionally, a wide range of dietary factors, including meat and vegetable consumption, were associated with reduced kidney aging (Figure 7A).

We examined the relationship between intake of drinks and organ aging, comparing high (≥75^th^ percentile) consumption to moderate (25^th^-75^th^ percentile consumption) and low (≤25% percentile) to moderate consumption. Here, we found that high water intake was modestly associated with increased brain aging, consistent with clinical literature suggesting excess fluid intake and thirst can reflect underlying physiological dysregulation ^43^. We additionally find that high versus moderate intake of coffee, tea, and water is linked to reduced kidney aging (Figure 7B). We emphasize, however, that the magnitude of these associations is small and results should not be interpreted causally (Figure 7B).

Smoking and alcohol frequency were both strongly associated with accelerated aging across most organs, indicating potential systemic effects in accelerating aging. However, smoking showed weak associations with the hips and knees, and higher alcohol frequency was associated with modestly reduced aging in the kidneys and pancreas (Figure 7C-D).

Waist-to-hip ratio, a marker of obesity, was strongly associated with accelerated aging across a wide range of organs, suggesting, like smoking and alcohol, a potential system-wide pro-aging effect. Of these organs, pancreas aging was most strongly associated with waist-to-hip ratio, followed by the heart atria, kidneys, and various brain regions. Intriguingly, waist-to-hip ratio was associated with modestly decreased aging in the knees (Figure 7E).

Physical activity levels showed divergent associations across organs––increased aging in the hips and knees, potentially a function of exercise-induced wear and tear, and decreased aging in torso organs, such as the pancreas and kidney, as well as in the two full-body models (Figure 7F).

Notably, a sense of life meaning was associated with reduced brain aging, with the strongest region-specific links observed in the cerebrum and ventral diencephalon (Figure 7G). This suggests that a sense of purpose may have a protective effect on brain aging, concentrated in specific regions. Sense of happiness was likewise associated with reduced brain aging (Figure S6C). We did not observe any major associations between childhood abuse or lifetime depression and organ-specific aging (Figure S6D-E).

Examining sleep duration revealed distinct relationships for abnormally long (>9 hours) and abnormally short (<7 hours) sleep durations compared to normal durations (7-9 hours).

Abnormally long sleep was linked to increased brain aging, consistent with literature on prolonged sleep associating with dementia risk ^44,45^. Conversely, abnormally short sleep was associated primarily with increased cardiac and skeletal aging (Figure S6A-B).

We also examined several eye surgeries and found, remarkably, that cataract surgery may reverse features of eye aging captured by our model (Figure 7H). To analyze this further, we examined the association between left retina fundus aging and cataract surgery of the left eye and right eye separately. Strikingly, we found that only left eye (but not right eye) cataract surgery was associated with reduced retina fundus age of the left eye (Figure S6H-I), pointing toward the existence of a causal relationship between cataract surgery and reversal of eye aging markers. Refractive laser eye surgery and glaucoma surgery displayed no relationships with organ aging (Figure S6F-G).

Given the complex interrelationships and covariation among lifestyle factors, we performed elastic net regression analysis to identify the most salient predictors of organ aging while accounting for covariation among variables. This analysis revealed notable patterns across organs. Waist-to-hip ratio, a marker of central obesity and metabolic health, emerged as the strongest predictor of increased organ aging across a range of organs, with coefficient magnitudes substantially larger than other lifestyle factors (Figure S7D), suggesting that overall metabolic health and central adiposity are far more predictive of organ aging patterns than specific dietary choices or lifestyle behaviors.

## Discussion

Our study presents a new paradigm to noninvasively assess human aging across a comprehensive set of organs, tissues, and anatomical regions at sub-organ spatial resolution. Through the combination of multi-modal imaging, segmentation algorithms, and deep learning networks applied to high-dimensional imaging data, we provide, to our knowledge, the most spatially detailed profiling of organ-specific aging to date in a total of over 130K people. Our approach reveals new insights into the heterogeneity of aging both within and across individuals, the relationship between organ aging and disease, and the potential influence of modifiable lifestyle factors on the aging process at organ resolution.

Our research builds on a rich literature of image-based analysis of biological aging, including studies assessing aging via brain MRIs ^13,46–48^, abdominal MRIs ^14^, cardiac MRIs ^49,50^, retinal imaging ^51,52^, and DEXA scans ^53^, as well as recent work tracking organ aging from image-derived phenotypes (IDPs) ^8^. Building on this foundation, our study comprehensively integrates five imaging modalities and applies state-of-the-art segmentation algorithms to these images. This approach enables the isolation and analysis of organ-specific aging trajectories at unprecedented scale and resolution.

The advantage of our organ-specific approach is particularly evident in the context of disease risk prediction. Models trained exclusively on segmented pancreas features demonstrated robust associations with incident type 1 and type 2 diabetes, and kidney-specific models were most predictive of future kidney disease development. However, these organ-disease associations were weaker in the whole-torso model, demonstrating that organ segmentation unmasks critical organ-specific biological signals that are otherwise diluted in whole-image analysis. Importantly, our image-based approach reflects only the cellular aging processes in a particular organ that lead to phenotypic changes, without capturing non-functional cellular alterations. Moreover, in contrast to IDP-based methods, which rely on a limited number of predefined measurements per organ, our deep learning approach leverages the full high-dimensional information contained in medical images to capture complex morphological aging patterns at sub-organ resolution. Requiring no tissue collection or sample processing, our approach facilitates rapid quantification of organ aging, making it highly conducive to frequent and longitudinal assessment.

The correspondence between our image-based organ aging metrics and organ-specific circulating biomarkers represents a fundamental validation that non-invasive imaging can serve as a window into the complex physiological and molecular processes underlying organ aging. It also highlights that structural and functional aging processes are inherently connected, suggesting imaging may serve as a powerful surrogate for invasive functional assessments and tissue biopsies in clinical practice.

Our attention maps consistently highlighted biologically meaningful regions and revealed several noteworthy patterns, particularly for the skeletal aging models. The full-body skeleton model highlighted the skull and torso as being most relevant for capturing aging. These areas encompass vital organs crucial for survival, indicating the model captures features with significant physiological impact. In this model, attention was concentrated on the spine. In direct alignment with this, the spine is particularly susceptible to structural damage with age due to its complex structure and manifold functions, which contribute to its pronounced age-related frailty compared to less mobile bones ^54^.

In the hips and knees, our models focused particularly strongly on the joints. Consistent with this, the interface between bones––rather than the bones themselves––is significantly more susceptible to age-related wear and damage due to its dual functions of bearing weight and facilitating movement ^55^. Altogether, our skeletal model attention maps suggest that age-related morphological changes are most evident in regions that both undergo biomechanical stress and enable movement, indicating these areas may be the most promising targets to mitigate age-related musculoskeletal dysfunction.

We found that organ aging strongly predicted corresponding organ disease across a wide range of cases. Beyond these organ-disease-corresponding relationships, we observed several intriguing inter-organ relationships. For instance, kidney aging was robustly associated with heart failure, which is consistent with well-established evidence that kidney disease drives cardiac dysfunction ^56–58^. Additionally, pancreas aging predicted the risk of developing kidney disease ^59^, consistent with established evidence that kidney disease is a well-known complication of pancreas-associated type 2 diabetes ^59^.

While our associative analyses cannot establish causality, these findings, consistent with established literature on organ dysfunction and disease relationships, begin to characterize the complex networks connecting organ aging to organ disease.

We found that aging of the brain, and particularly the cerebrum, was among the strongest predictors of organism-wide aging outcomes. Consistent with our findings, organ aging models derived from plasma proteins also identified brain-specific proteins as strong predictors of increased mortality risk in humans ^60^. Moreover, a youthful brain profile was associated with enhanced longevity ^60^. Together, these lines of evidence suggest that the brain may serve as a central regulator of systemic aging and that interventions targeting brain aging may have the most potential to impact organismal aging and lifespan.

The stark contrast we discovered between cerebral and cerebellar aging was particularly striking. While cerebral aging was robustly predictive of mortality and morbidity, cerebellar aging showed minimal associations with both outcomes. This suggests the cerebrum and cerebellum may age in fundamentally different ways. Our findings align with epigenetic studies showing that the cerebellum has a slower aging rate compared to other brain regions ^61^.

However, recent work has also observed age-related gray matter volume loss in specific cerebellar clusters predominantly involved in non-motor cognitive functions ^61^. This suggests that while cerebellar aging may impact specific functional domains, it does not significantly contribute to overall organismal aging at the level of lifespan and major disease development.

Investigating Alzheimer’s disease, we found that only a subset of brain regions, including the cerebral cortex and several subcortical structures like the hippocampus, were predictive of disease onset. These subcortical structures play crucial roles in memory and emotional regulation, which are impaired in early Alzheimer’s disease ^62,63^. Our analysis suggests that Alzheimer’s disease emerges from, and may be caused by, aging of only certain brain regions, rather than following from a global pattern of brain aging.

Analysis of other diseases revealed highly distinct patterns in their relationship to organ aging. Hip arthritis and age-related macular degeneration (AMD) were associated largely with hip aging and retina aging, respectively, suggesting they emerge mostly from region-specific aging processes. In contrast, type 2 diabetes and kidney disease, while also linked to aging of the pancreas and kidney, respectively, were associated with aging of a much broader set of organs, suggesting an association with systemic rather than region-specific aging. Consistent with this, previous studies have shown that both type 2 diabetes and kidney disease considerably affect systemic health ^64–67^.

Our analysis of lifestyle factors revealed complex, organ-specific associations that challenge simplistic paradigms of healthy versus unhealthy behaviors. However, we emphasize that these findings must be interpreted with considerable caution given the observational nature of our study design, which cannot establish causality.

Among dietary factors, we found meat consumption correlated with reduced skeletal aging. While this aligns with established nutritional literature suggesting adequate protein intake is crucial for maintaining bone density and counteracting frailty in aging populations, this should be balanced against established associations between excess protein intake and increased risk of age-related diseases ^68^. The association between intake of various food types with reduced kidney aging limits any definitive interpretations given the lack of food-type specificity.

Our findings regarding brain aging and mental well-being were particularly striking, with both sense of purpose and happiness displaying robust links to reduced aging in specific brain regions, particularly the cerebrum and ventral diencephalon. This aligns with previous studies demonstrating a link between a higher sense of purpose and reduced mortality risk ^69^, including studies suggesting a causal relationship between mental well-being and longevity ^70^. The region-specificity of these events provides insight into potential biological mediators between psychological well-being and increased longevity.

Together, our results highlight the importance of reconceptualizing aging as a series of parallel processes simultaneously occurring across different organs and tissues rather than as a uniform phenomenon. These parallel processes may interact through organ aging networks, in which aging of one system affects the aging trajectory of other organs through complex feedback loops. Alternatively, these processes may occur largely in isolation, such as in the retina or joints. Crucially, aging of different systems may have vastly different implications for disease risk. Some diseases may develop from localized aging (as in AMD), while others may emerge from multi-system aging processes (as in type 2 diabetes). Similarly, the variability in organ disease development with age likely reflects variability in organ aging patterns preceding disease onset.

Likewise, aging of different systems may have dramatically different effects on mortality risk and systemic aging, with aging of some organs, like the cerebrum, potentially raising mortality risk, while aging of other organs, e.g., the cerebellum, may not be as closely associated with mortality risk. Similarly, organs across the body may have hierarchical relationships with systemic aging. For instance, as our results suggest, pancreas aging may be more critical than kidney aging, which may be, in turn, more important than liver aging. These hierarchical relationships may reflect the varying extent to which deleterious changes manifest in these organs with age.

Our results indicate that existing lifestyle interventions known to impact health may affect these parallel aging processes differently or have an effect only in a particular organ system. This suggests that developing interventions targeted to specific organs and systems may be more feasible than developing an intervention that can simultaneously target aging across each of these disparate parallel processes. Our results suggest that interventions targeted to the organs most relevant to organismal aging, e.g., potentially the cerebrum or pancreas, may have an outsized influence in extending healthy lifespan, whereas interventions targeted to other organs, like the cerebellum or liver, may yield only minor effects.

In the future, several steps could further enhance the scope and capabilities of our approach. Improvements in automated segmentation algorithms will likely enable the analysis of a much broader range of organs and tissues from bulk-level images. For MRIs, recent technological progress now enables imaging at mesoscale resolutions ^71^, which, combined with segmentation improvements, may ultimately facilitate non-invasive profiling of aging across thousands of anatomical regions at near-cellular resolution. This could revolutionize our understanding of human aging. Additionally, future studies could better characterize the causal relationships between aging of different organs through approaches such as Mendelian randomization.

Our study establishes a powerful framework for assessing human aging at organ resolution through non-invasive imaging. Using widely available medical imaging and automated segmentation approaches, we reveal new insights into the heterogeneity of aging across organs and individuals. The ubiquity of organ-specific extreme aging––affecting nearly one in six people––highlights the critical importance of moving from conventional, single-metric measures of aging to organ-specific assessment.

Integrating this approach into clinical practice––using medical images that are already routinely generated––could transform how aging is monitored and managed. By providing a framework to easily assess organ aging, this approach creates the opportunity to develop and apply proactive interventions targeting accelerated organ-specific aging before the development of organ disease. Together, these advances represent a significant step in understanding, monitoring, and managing the aging process to improve human health as we age.

### Limitations of the study

Several limitations to this study should be noted. First, while our approach leverages validated segmentation algorithms, these tools may generate imperfect segmentations. Although we filtered out images with potential segmentation errors based on outlier dimensions (see Methods), the scale of our analysis made manual review of each segmentation infeasible.

However, while previous studies have shown that segmentation accuracy is associated with age, this effect is minimal ^21^. Second, we emphasize that our analyses are associative and cannot establish causality. Our results may be subject to reverse causality and, despite adjusting for relevant confounding factors, may also be influenced by unmeasured confounding factors. Additionally, the UK Biobank is notably limited in terms of population diversity, consisting mostly of people of Northern European ethnicity ^72^. Future validation of our approach and findings in more ethnically diverse cohorts will be essential to ensure their generalizability.

## Methods

### Ethics

This study was approved by the Mass General Brigham Institutional Review Board.

### UK Biobank cohort

The UK Biobank is a biomedical research database with a cohort of approximately 500,000 individuals in the United Kingdom. From these individuals, comprehensive data is collected on lifestyle factors, genomics, plasma proteomics and metabolomics, health records, imaging, and physiological measures. This study analyzed a subset of the UKB cohort with imaging data available (134K subjects). The number of total images available for each image type is provided in Table S2A and demographic information for the UKB imaging cohort analyzed in this study is presented in Table S2B.

### Image generation

The UK Biobank has collected multiple imaging modalities from participants, including baseline retinal images and additional imaging as part of the UK Biobank imaging study ^73^. Our study utilized T1 brain MRIs, torso MRIs, cardiac MRIs, DEXA images, and retinal fundus scans.

#### Brain T1-weighted MRI

T1-weighted structural brain images were captured with a 3 Tesla Siemens MAGNETOM Skyra scanner using a 32-channel head coil and VD13 software. Images underwent an automated processing and quality control pipeline using FSL and FreeSurfer ^73^.

#### Cardiac MRI

Cardiac MRIs were acquired with the Siemens 1.5 Tesla MAGNETOM Aera scanner with VD13A software. The protocol captured cardiac structure and function through long-axis cine imaging, short-axis cine imaging, and transverse aortic cine imaging. Further details are described in Petersen et al., 2016 ^74^.

#### Torso MRI

Torso MRIs (also referred to as abdominal MRIs in other publications) were performed with the same Siemens 1.5 Tesla MAGNETOM Aera scanner. The protocol included multiple sequences targeting the liver, pancreas, and entire torso, which included the hips and upper legs ^73^.

#### DEXA

DEXA scans were taken with the GE-Lunar iDXA instrument. Imaging included two total-body images, one highlighting flesh and one highlighting the skeletal system, and specific images of the left and right hip and knees and two views of the spine: anterior-posterior (AP) view and lateral-vertebral assessment (LVA) view ^73^.

#### Retina Fundus

Retinal fundus images were captured with the Topcon 3D OCT1000 Mark II ophthalmic camera, capturing 45° field-of-view images centered on the optic disc and macula ^75^.

### Image processing and segmentation

#### Torso MRI

Torso MRIs were segmented using software developed and validated in Kart et al., 2021 and Kart et al., 2022 ^20,21^. We applied this software in 4-channel mode to segment the left and right kidneys, liver, pancreas, and spleen, generating masks for each organ. To enhance performance in predicting age, we combined the left and right kidney masks into a single kidney mask.

For each organ, we applied the corresponding segmentation mask to the in-phase torso MRI images and set values outside the mask to zero. The resulting organ-specific volumes were cropped to the maximum dimensions of the segmented volume and resized to the median dimensions across all segmented volumes. We then z-score normalized the intensity values. Any images with corresponding segmentation masks with a volume of zero were excluded from analysis.

Images were further filtered based on segmented dimensions: any image with outlier dimensions (sagittal, coronal, or axial) was excluded to omit images with potential segmentation errors. Outliers were defined as values above Q3 + 1.5 * IQR or below Q1 - 1.5 * IQR, where Q3 is the third quartile, Q1 is the first quartile, and IQR is the interquartile range of a particular dimension.

For the full torso volume, we extracted 25 evenly spaced slices along the sagittal axis to reduce memory usage during subsequent model training.

#### Brain MRI

We began with T1-weighted brain MRIs that were defaced and converted to MNI152 space. To segment these images, we applied SynthSeg ^19^, which yielded masks for 32 brain regions. As with the torso MRIs, for each brain region, we applied the corresponding segmentation mask to the original T1 brain MRI image, cropped the resulting volumes to the maximum nonzero dimensions, resized images to the median segmented dimension, and z-score normalized the intensity values. Images whose corresponding segmentation masks had a volume of zero or whose segmented dimensions were considered outliers (as defined above) were filtered out.

#### Cardiac MRI

We segmented the cardiac MRIs using software developed and validated in Bai et al., 2018 ^17,18^. Specifically, we segmented the ventricles from the short axis cine images, the atria from the long axis 4 chamber images, and the aorta (ascending and descending) from the transverse aortic images.

From the long axis and short axis cines, we selected images corresponding to end-systole to normalize the cardiac cycle across images. To convert each image to a 3-channel format compatible with 2D CNNs, we applied the following steps: for the short axis images (which were 3-dimensional), we took the three middle slices. For the long axis images (which were 2-dimensional), we triplicated the images to yield 3 channels. For the aorta images (which were 2-dimensional cines), we isolated and triplicated the first frame. As with the brain and torso MRIs, we applied the corresponding mask to each image type; however, here we dilated the mask slightly to ensure the heart wall and aortic wall were included in the segmented image. Then, we cropped the segmented images based on the maximum nonzero dimension and resized them to the median dimensions. If either the width or height of the median dimensions were less than 75 pixels, we resized images on that dimension to 75 pixels to make them compatible with the 2D CNN we developed. Finally, we z-score normalized the intensity values. As before, images whose segmentation masks had a volume of zero or whose segmented dimensions were considered outliers (as defined above) were excluded from further analysis.

#### DEXA

For the DEXA images, we utilized the following images: region-specific images, including the left hip, right hip, left knee, right knee, AP spine, LVA spine; and two full-body images, one highlighting flesh (called *full-body flesh* here), and one highlighting the skeleton (called *full-body skeleton*). Each greyscale image was resized to the dimensions (299 x 299) to reduce memory pressure and triplicated to create 3-channel images compatible with our 2D CNN before z-score normalization.

#### Retina fundus

We first cropped each image to match the radius of the fundus circle. Next, we applied quality filters to exclude images that did not meet specific criteria. Images were omitted from analysis if the fundus circle could not be identified, if they were overexposed (maximum brightness in non-red channels >165), or if they lacked sufficient red intensity (red intensity >20), defined as follows:

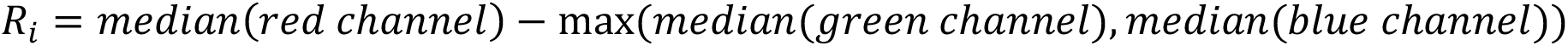

These metrics were based on previously developed methods ^76^. Then, images were resized to 299 x 299 x 3 dimensions and z-score normalized.

### Machine learning models

We constructed two convolutional neural networks (CNNs) to generate image-based age estimates: one designed for 3D images and one for 2D images.

### 3D convolutional neural network

For 3D images, we built a ResNet-based CNN to predict age using both 3D images and sex as inputs. Our base model begins with a 3D convolutional layer, which is followed by batch normalization, Swish activation, and max pooling. We then added a series of four custom residual blocks, each with three 3D convolutional layers: two connected to each other and one a skip connection. Each residual block contains batch normalization and activation layers. After the ResNet base model, we applied a global average pooling layer to reduce dimensionality and concatenated the output with a 16-node side neural network to incorporate sex as an input, which enables the model to interpret images differently depending on the sex of the participant. Following this, we added a dropout layer for regularization, a fully connected layer with ReLU activation and L2 regularization, a second dropout layer, and a final fully connected layer with a single node and linear activation to predict age (Figure S2A).

### 2D convolutional neural network

For the 2D images, we constructed a CNN to predict age using 3-channel images and sex as inputs. This model utilizes the InceptionV3 architecture as a base, followed by a global average pooling layer to reduce dimensionality. The model also incorporates a 16-node side neural network to input sex. Following these layers, we added a dropout layer, a fully connected layer with L2 regularization and ReLU activation, a second dropout layer, and a final fully connected layer with one node and linear activation to output age (Figure S2B).

### Model training

#### Training details

To train our models, we utilized the Adam optimizer and Huber loss function, an intermediate between mean absolute error and mean squared error loss functions, which we found improved performance for some image types. To prevent overshooting, we implemented a learning rate scheduler that reduces the learning rate by a factor of 0.5 if the model shows no improvement over five epochs. We also applied early stopping with a patience of 15 epochs during hyperparameter optimization and 25 epochs during final model training to prevent overfitting and reduce training time. Models were trained for a maximum of 250 epochs. We scaled the target variable, chronological age, by a factor of 0.01, which we found improved training stability. We implemented our models using the TensorFlow (version 2.14) and Keras (version 2.14) libraries and NVIDIA Tesla V100 GPUs for all models except the cardiac MRI models, which required us to utilize the UKB-RAP platform (Tensorflow version 2.11, Keras version 2.11) due to the UK Biobank’s transition to the platform.

#### Data augmentation

For the 2D CNN, we applied data augmentation. Specifically, we introduced random rotations and translations, the extent of which was controlled by a hyperparameter (details provided below).

#### Cross-validation and cohort selection

To capture features of aging while minimizing the influence of major confounding factors and disease, we selected a generally healthy cohort in which to train our models. Participants were filtered based on several criteria including smoking history, current pregnancy status, self-reported health, number of past diseases, obesity, and other factors listed in Table S1A. Metadata features for the training cohort and full imaging cohort are plotted in Figure S1D and Figure S1F.

We sought to generate cross-validated, out-of-fold age predictions for as many participants as possible while keeping training times reasonable. The training times required to perform hyperparameter optimization made it impractical to perform hyperparameter optimization for each cross-validation fold. Instead, we randomly selected 25% of the generally healthy cohort for hyperparameter tuning. This subset was excluded from further analysis to ensure that optimized hyperparameters were applied only to images with which they were not specifically optimized. The optimized hyperparameters were then applied to generate cross-validated age predictions in the remaining generally healthy cohort by training on 80% of the data and predicting on the final 20% across five consecutive folds. Finally, we trained a model on all generally healthy participants who were not part of the hyperparameter optimization set, then used this model to generate age predictions for participants outside the generally healthy group.

To further reduce training time, we applied optimized hyperparameters across similar images. For brain MRI models, hyperparameters were optimized on whole-brain and brainstem images, and brainstem-derived parameters were applied to other segmented brain regions. For DEXA images, optimization was performed on the AP spine and full-body skeleton images. Then, AP spine hyperparameters were applied to other skeletal components and full-body skeleton hyperparameters were applied to the full-body flesh image. For cardiac and torso MRI models, we optimized each segmented image type separately to improve performance.

Hyperparameters for pancreas images were based on those optimized for liver images, which yielded better accuracy.

#### Hyperparameter optimization

Hyperparameter tuning was conducted using Optuna ^77^ for 60 trials, optimizing parameters including the L2 regularization weight, dropout rate, initial learning rate, training batch size, and the number of nodes in the penultimate fully connected layer. For the 3-channel CNN, we also optimized the strength of augmentations. For the 2D models, we tested both 32 and 64 training set batch sizes; however, for the heart atria models, a batch size of 32 resulted in training errors, so we specified a batch size of 64. The full set of hyperparameters and ranges are provided in Table S1B.

#### Model selection

We retained only models with a Pearson correlation coefficient across all out-of-fold participants of at least 0.4 between predicted and chronological age before bias correction (detailed below). The total number of images for which out-of-fold age estimates were generated for each model is listed in Table S3. We compare the performance of our models to plasma protein-based models ^24^ in Figure S1C and note comparable performance.

### Attention maps

To identify image regions most relevant for age prediction, we generated attention maps using the Grad-CAM method ^35^, which calculates the gradient of the output with respect to features in the final convolutional layer. For each image type, attention maps were computed using models that had not been trained on the respective images.

For DEXA, retinal images, and unsegmented brain and torso MRIs, we applied Grad-CAM to the last convolutional layer. However, for other image types, the smaller input image sizes made the final convolutional layer too small to generate informative features. To address this, we qualitatively selected earlier convolutional layers that provided interpretable features: the 6th-to-last layer for segmented torso images, the 9th-to-last layer for segmented brain images, and the 90th-to-last layer for cardiac MRI images.

### Age deviation bias correction

We observed a significant bias in age deviation (predicted age minus chronological age) across different age groups: younger individuals were consistently predicted to be older than their chronological age, while older individuals were predicted to be younger. This bias is a well-documented statistical artifact that may confound analysis ^14^. To correct this bias, we applied a linear regression adjustment before further analysis. This adjustment followed the Cole’s method ^78^, which is detailed here: ^79^.

### Health outcomes definitions

Cancer outcomes were identified using the cancer registry, dates of death from the death registry, and non-cancer diseases based on the first occurrences category in the UK Biobank. The time to a health outcome event was calculated as the time from the imaging visit to the event (disease diagnosis or death), and individuals who did not experience the event were considered censored. Censoring dates were based on those recommended by the UK Biobank at the time of this study: for death, November 30, 2022; for non-cancer disease, May 31, 2022; for cancer in Wales, December 31, 2016; for cancer outside Wales, December 31, 2020. We inferred whether a participant lived in Wales based on the assessment center location of their first visit. For a given disease, if death occurred before the censoring date, the censoring date was updated to the date of death.

We defined morbidity, or “healthspan end,” as the date of the first occurrence of either death or a major disease diagnosis. Major diseases included in this definition were cancer, dementia, Parkinson’s disease, motor neuron disease, stroke, ischemic heart disease, heart failure, chronic obstructive pulmonary disease (COPD), type 1 diabetes, type 2 diabetes, liver disease, and kidney disease. We used the earliest possible censoring date for morbidity, which was the censoring date for cancer. Additional details and criteria for each disease are provided in Table S5.

### Quantification and statistical analysis

All statistical analyses were performed in R (version 4.3.0). A *P*-value threshold of 0.05 was used for determining significance. *P*-values were adjusted for multiple testing with the Benjamini-Hochberg false discovery rate correction wherever appropriate.

### Incident health outcome associations

To assess the association between organ age predictions and the risk of incident health outcomes (disease and mortality), we applied a Cox proportional hazards model to compute hazard ratios between z-score normalized organ age and health outcome risk while adjusting for chronological age and sex. This was performed with the coxph function from the survival package in R ^80^. For each disease, we excluded individuals already diagnosed with the disease at baseline or diagnosed within a year of the imaging visit. We selected 25 of the most common and organ-specific diseases to analyze as well as all-cause mortality and age-related morbidity, all of which are listed in Table S5.

### Local versus remote organ predictive analysis

To investigate whether anatomically remote organs’ aging features improved predictive value for organ-disease prediction beyond local organs’ aging processes, we compared the predictive performance of anatomically relevant “local” organs versus the same local organs combined with anatomically remote organs. We conducted this analysis across six major diseases affecting organs captured in our models during the UKB imaging visit: Alzheimer’s disease (local: whole brain), type 2 diabetes (local: pancreas), hip osteoarthritis (local: left hip), knee osteoarthritis (local: left knee), kidney disease (local: kidneys), and ischemic heart disease (local: heart atria). For each disease, we fitted Cox proportional hazard models using Survival’s coxph function with the local organ’s aging score, chronological age, and sex, with and without remote organ ages, assessing improvement with Harrel’s C-statistic and quantifying statistical significance with the likelihood ratio test. As before, for each disease, we excluded individuals already diagnosed with the disease at baseline or diagnosed within a year of the imaging visit.

### Extreme agers analysis

To assess the frequency of organ-specific extreme aging, we analyzed a subset of distinct organs all captured during the imaging visits: left hip, right hip, left knee, right knee, AP spine, kidneys, liver, pancreas, atria, aorta, and brain. Retinal imaging was excluded as it was not part of the imaging visit, and thus, there were very few individuals with retinal imaging in addition to other imaging modalities (and other organ age estimates) for the same visit. We analyzed only images from the first imaging visit to avoid including duplicate participants. For each organ, we calculated the proportion of participants exhibiting an age deviation of at least 2 standard deviations above the mean in that organ and no other organs for the same individual, building on previous analyses of organ-specific extreme aging ^5^. The age distribution of extreme agers compared to non-extreme agers is plotted in Figure S1G.

### Molecular and physiological biomarker associations

For each organ aging estimate, the temporally closest biomarker measurement within a 10-year window was selected from the same UK Biobank participant. This matching was performed separately for each biomarker subset, including plasma proteomics (Olink platform), plasma metabolomics (Nightingale Health NMR platform), blood biochemistry, and physiological measures. Residuals were calculated for organ age predictions and each biomarker by fitting linear models that included chronological age as a covariate. Pairwise partial Spearman correlations were then calculated between the age-adjusted organ age deviation values and individual biomarkers. The results for the molecular associations (including blood biochemistry, plasma proteomics, and plasma metabolomics) are provided in Table S3.

### Pathway enrichment analysis

To gain insights into the biological pathways underlying organ aging patterns, we conducted gene set enrichment analysis (GSEA) ^30^ of plasma proteins associated with each organ’s aging process. We focused our analysis on a subset of major representative organs (pancreas, kidney, liver, aorta, left hip, spine, left knee, brain, retina, heart atria).

For each organ, we ranked all plasma proteins by their partial Spearman correlation coefficient with organ age deviation, creating organ-specific ranked gene lists. We then performed GSEA using the ClusterProfiler package’s GSEA function ^81^ against the Molecular Signatures Database (MSigDB) Hallmark gene sets ^31^.

### Lifestyle and intervention associations

To explore the relationship between lifestyle factors, interventions, and organ-specific aging, we performed multivariate linear regression analyses with organ age deviation as the outcome variable and lifestyle factors as predictors. Ordinal categorical variables (e.g., physical activity level) were treated as continuous variables in the regression models.

We also conducted elastic net regression analysis predicting organ age deviation as a function of all lifestyle factors with low (<15%) missingness to identify the most salient predictive factors while accounting for covariation among lifestyle variables. This was implemented with glmnet ^82^ in R with alpha = 0.5 and 10-fold cross-validation. This regularization approach enabled us to identify a minimal or parsimonious set of confounding covariates––waist-to-hip ratio and sex (Figure S7D).

For the multivariate linear regression analysis, we implemented three adjustment strategies to ensure the robustness of our results: (1) unadjusted models with no covariates, (2) parsimoniously adjusted models controlling for sex and waist-to-hip ratio, and (3) fully adjusted models controlling for chronological age, sex, physical activity level (IPAQ score), Townsend deprivation index, smoking status, and waist-to-hip ratio. When a covariate was the outcome variable of interest, it was omitted from the adjustment set.

To address potential confounding in dietary analyses, we excluded participants who reported making a major dietary change in the previous five years (field 1538) to reduce measurement error from inconsistent dietary patterns and mitigate reverse causation bias where individuals alter their diet in response to the onset of health conditions.

For beverage consumption analyses, we controlled for the intake of other beverages as covariates due to observed correlations between different beverage types (Figure S7A). We also stratified participants into consumption categories (low: ≤25th percentile, moderate: 25th-75th percentile, high: ≥75th percentile) to examine threshold effects.

For alcohol consumption analyses, we additionally controlled for mental health measures (general happiness, field 20458) and excluded participants who had changed their alcohol intake frequency in the previous 10 years (field 1628) to mitigate reverse causation from individuals who may have reduced consumption due to health conditions. For glaucoma surgery analyses, we additionally controlled for glaucoma diagnosis, and for smoking pack-years analyses, we included only past smokers.

We performed variance inflation factor analysis to confirm low collinearity among covariates used in fully adjusted models as well as all lifestyle factors together with low (<15%) missingness, reducing the risk of coefficient instability (Figure S7B-C).

## Resource availability

### Data and code availability

- To protect participant privacy, UK Biobank data is not publicly available; however, all *bona fide* researchers may apply for data access through the UK Biobank’s access management system.
- Code generated in this study will be made available upon publication.

## Data Availability

To protect participant privacy, UK Biobank data is not publicly available; however, all bona fide researchers may apply for data access through the UK Biobank's access management system.

## Acknowledgements

This research has been conducted using the UK Biobank Resource under Application Number 21988.

Reproduced by kind permission of UK Biobank ©.

## Authors’ contributions

A.E. conceived the study, performed analyses, and drafted the manuscript. D.G. developed the 3D convolutional neural network. A.M. analyzed and formatted the attention maps. A.D.Y. performed literature comparisons and analyses. A.T. performed the associations with blood biochemistry, plasma omics, and physiology measures. K.Y. developed the attention maps.

L.J.E.G. contributed to the comparison with the plasma protein models. C.M. assisted with the brain models’ development and analysis. V.N.G. supervised the study, guided analysis, edited and wrote the manuscript, and acquired funding. A.E., D.G., A.T., K.Y., and L.J.E.G. accessed UKB data. All authors contributed to manuscript preparation and approved the final version.

