## Supplementary Figures for "Anatomy of aging through organ-resolved multi-modal imaging and deep learning"

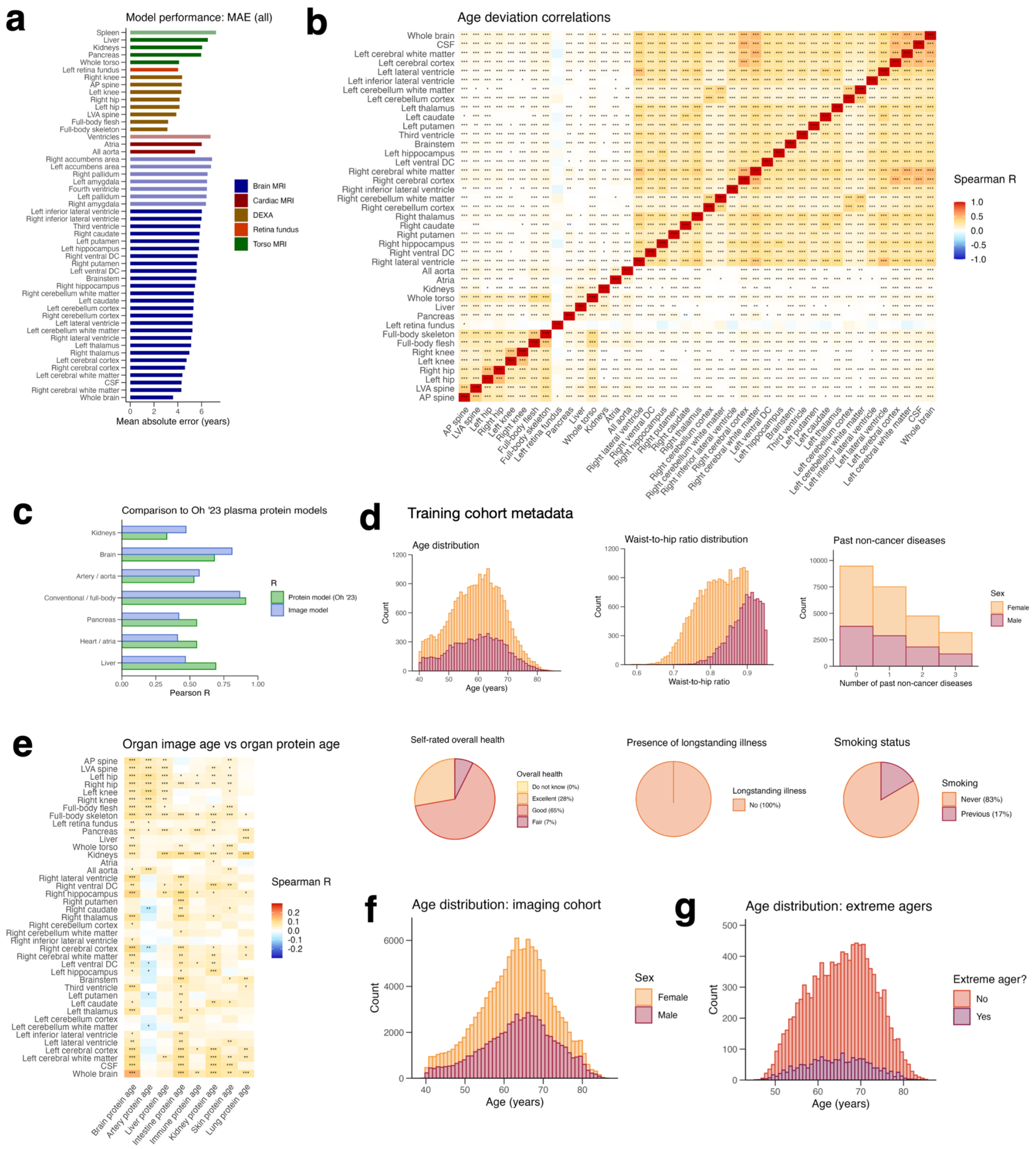


**Figure S1: Additional model performance and aging biomarker comparison. (a)** Model performance (mean absolute error) of each model in predicting age across all out-of-fold participants. Retained models exceeding the 0.4 Pearson R threshold are shaded more darkly. **(b)** Age deviation (predicted – chronological age) comparison across image models. **(c)** Model performance comparison to plasma protein-based models from Oh et al., 2023. **(d)** Metadata for generally healthy cohort used to train the image models. **(e)** Full comparison between image organ age deviation and organ protein age deviation. **(f)** Age distribution for UKB participants with imaging data. **(g)** Age distribution for extreme agers compared to non-extreme agers. Adjusted p-value < 0.001 = ***, adjusted p-value < 0.01 = **, adjusted p-value < 0.05 = *.

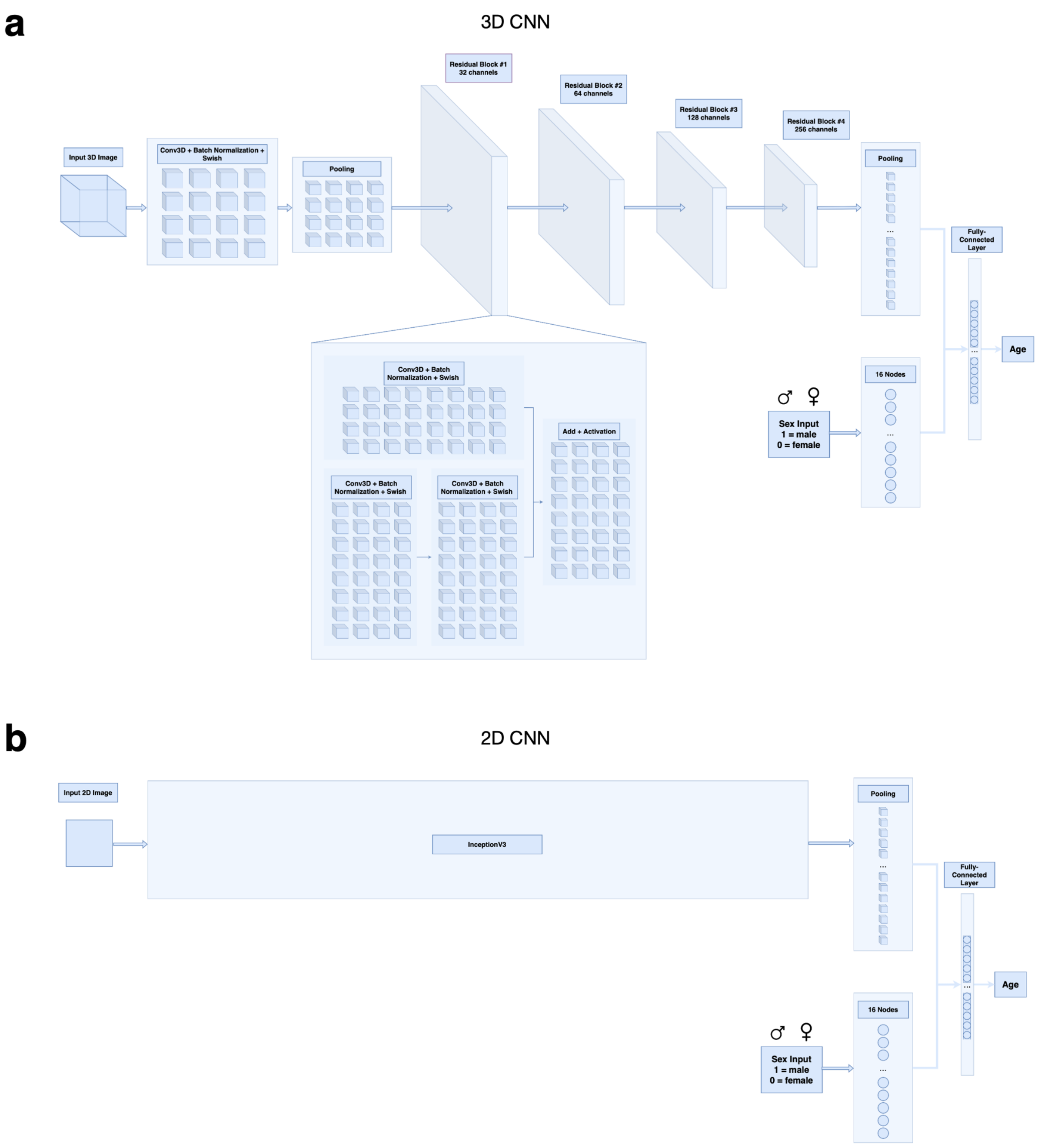


**Figure S2: Convolutional neural network model architecture.** Diagrams depicting the architecture of the 3D convolutional neural network **(a)** and the 2D convolutional neural network **(b)** that were developed for this study.

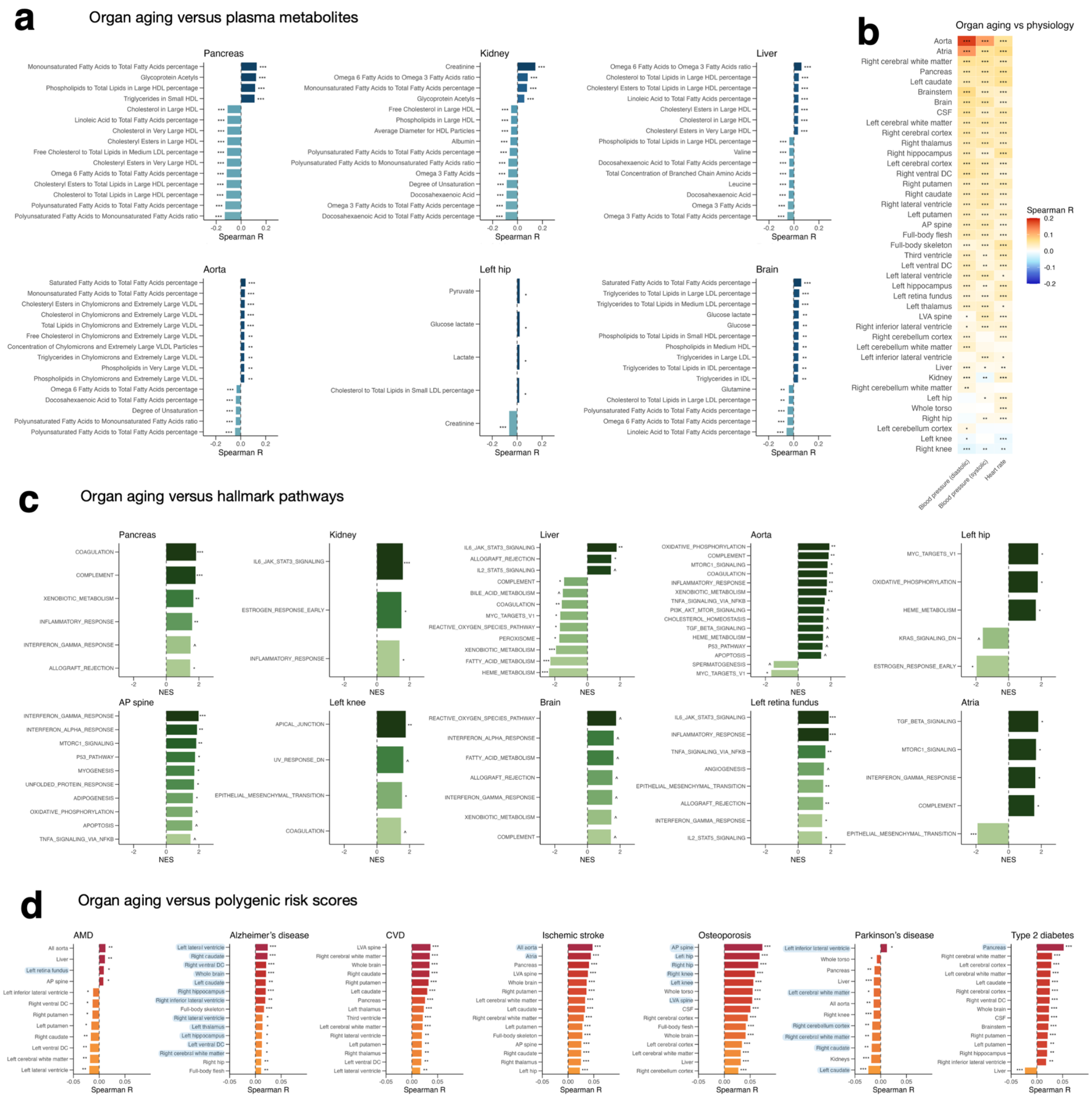


**Figure S3: Organ aging versus plasma metabolites, physiology, hallmark pathways, and polygenic risk scores. (a)** For a subset of major representative organs, correlation between organ age deviation and plasma metabolites (top 15 that were statistically significant shown here). **(b)** Correlation between organ age deviation and cardiac physiology (blood pressure and heart rate). **(c)** Gene set enrichment analysis results. For a subset of major representative organs, top 15 pathways (ranked by normalized enrichment score) with adjusted p-value < 0.1 are shown here. **(d)** Polygenic risk score analysis. For polygenic risk scores (PRS) computed for seven major age-related diseases, correlation between PRS and organ age deviation (top 15 organs that were statistically significant shown here). Adjusted p-value < 0.001 = ***, adjusted p-value < 0.01 = **, adjusted p-value < 0.05 = *, adjusted p-value < 0.1 = ^.

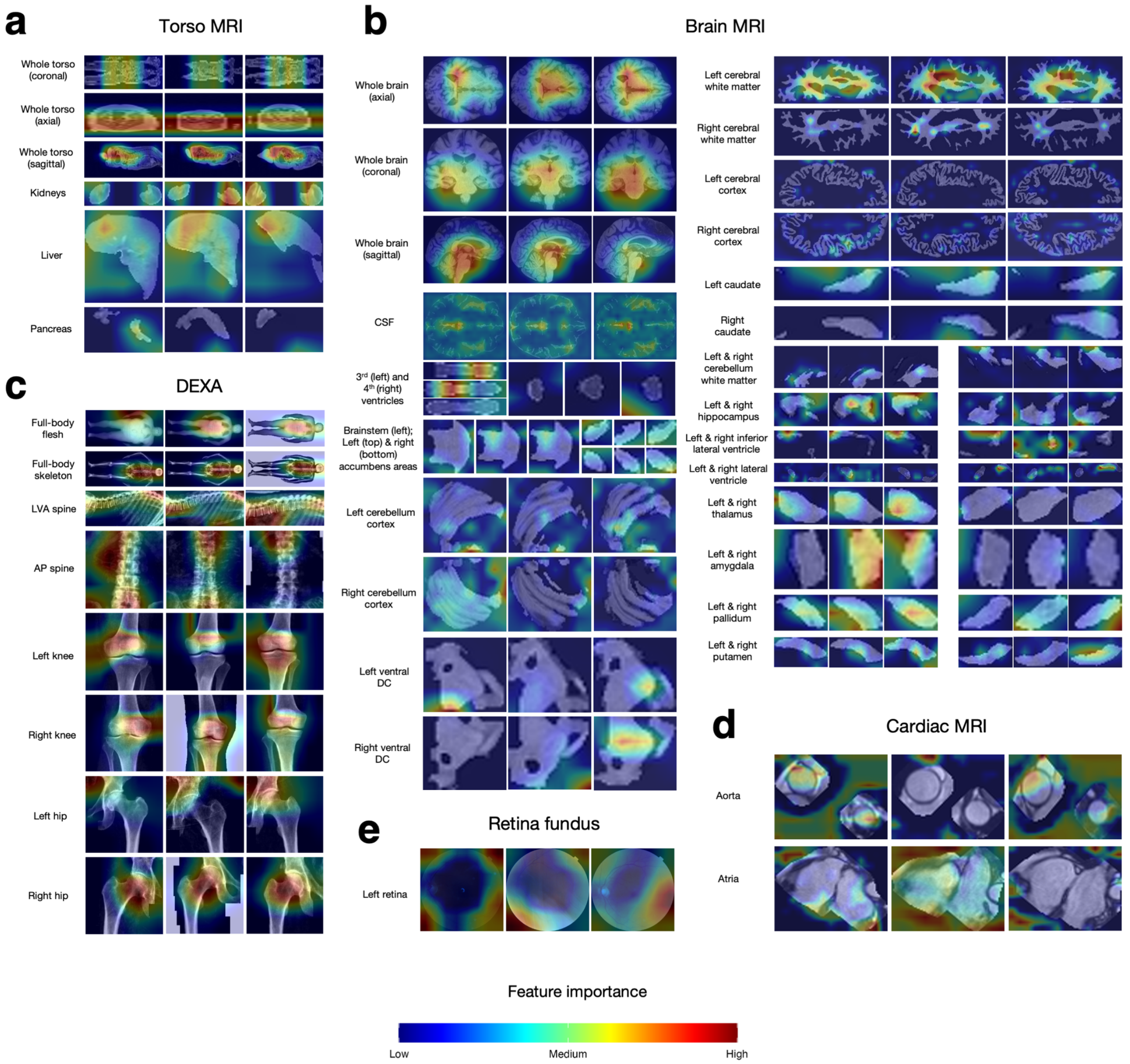


**Figure S4: Full set of attention maps.** Full set of attention maps of three individuals for the torso MRI models **(a)**, brain MRI model **(b)**, DEXA models **(c)**, cardiac MRI models **(d)**, and retina fundus model **(e)**. For the torso MRI models, all images are taken from the middle slice of the axial dimension except the whole torso model, which depicts the middle slice of the coronal, axial, and sagittal dimensions. Attention maps of three randomly selected people for each brain MRI model. For the brain MRI modes, all images are taken from the middle slice of the axial dimension except the whole brain model, which depicts the middle slice of the coronal, axial, and sagittal dimensions.

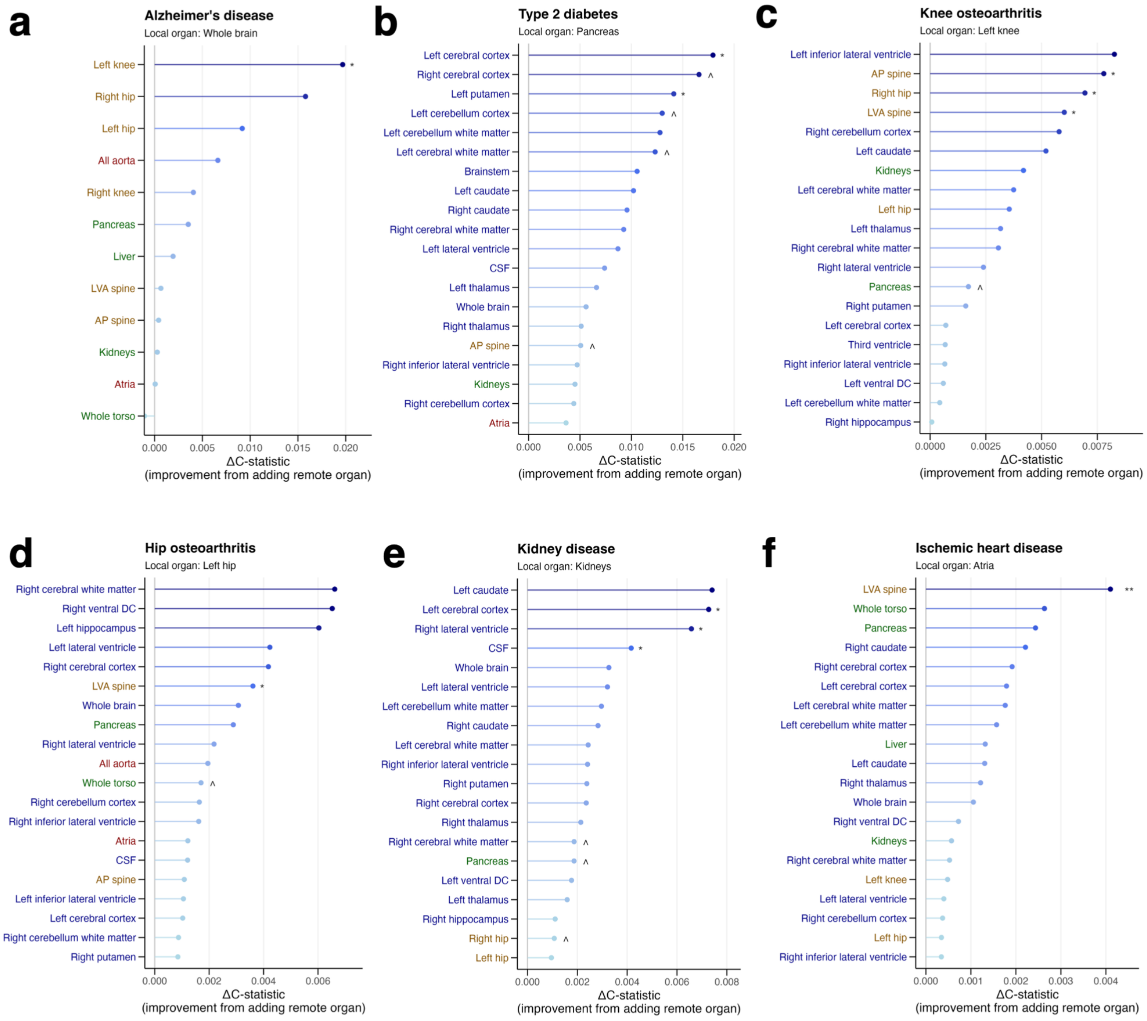


**Figure S5: Remote organ aging analysis.** Across six major age-related diseases, we quantify the predictive benefit of incorporating remote organs beyond organ-specific, local organs. Specifically, we analyze Alzheimer's disease **(a)**, type 2 diabetes **(b)**, knee osteoarthritis **(c)**, hip osteoarthritis **(d)**, kidney disease **(e)**, and ischemic heart disease **(f)**. The top 20 remote organs are shown here. Adjusted p-value < 0.001 = ***, adjusted p-value < 0.01 = **, adjusted p-value < 0.05 = *, adjusted p-value < 0.1 = ^.

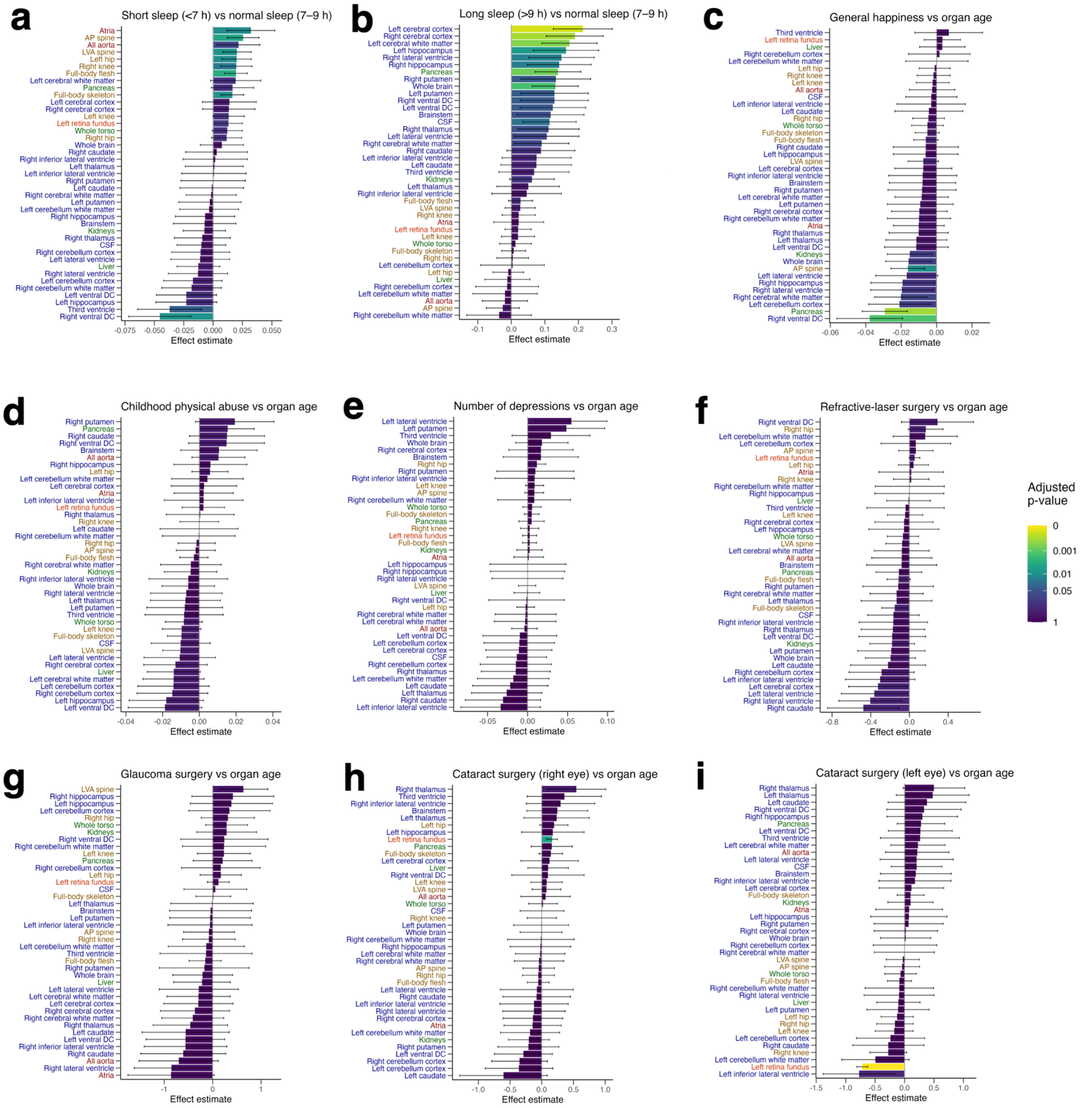


**Figure S6: Additional organ aging lifestyle and interventions analyses.** Relationship between organ aging and abnormally short sleep duration **(a)**, abnormally long sleep duration **(b)**, general happiness **(c)**, childhood physical abuse **(d)**, number of lifetime depressions **(e)**, refractive laser surgery **(f)**, glaucoma surgery **(g)**, right eye cataract surgery **(h)**, and left eye cataract surgery **(i)**.

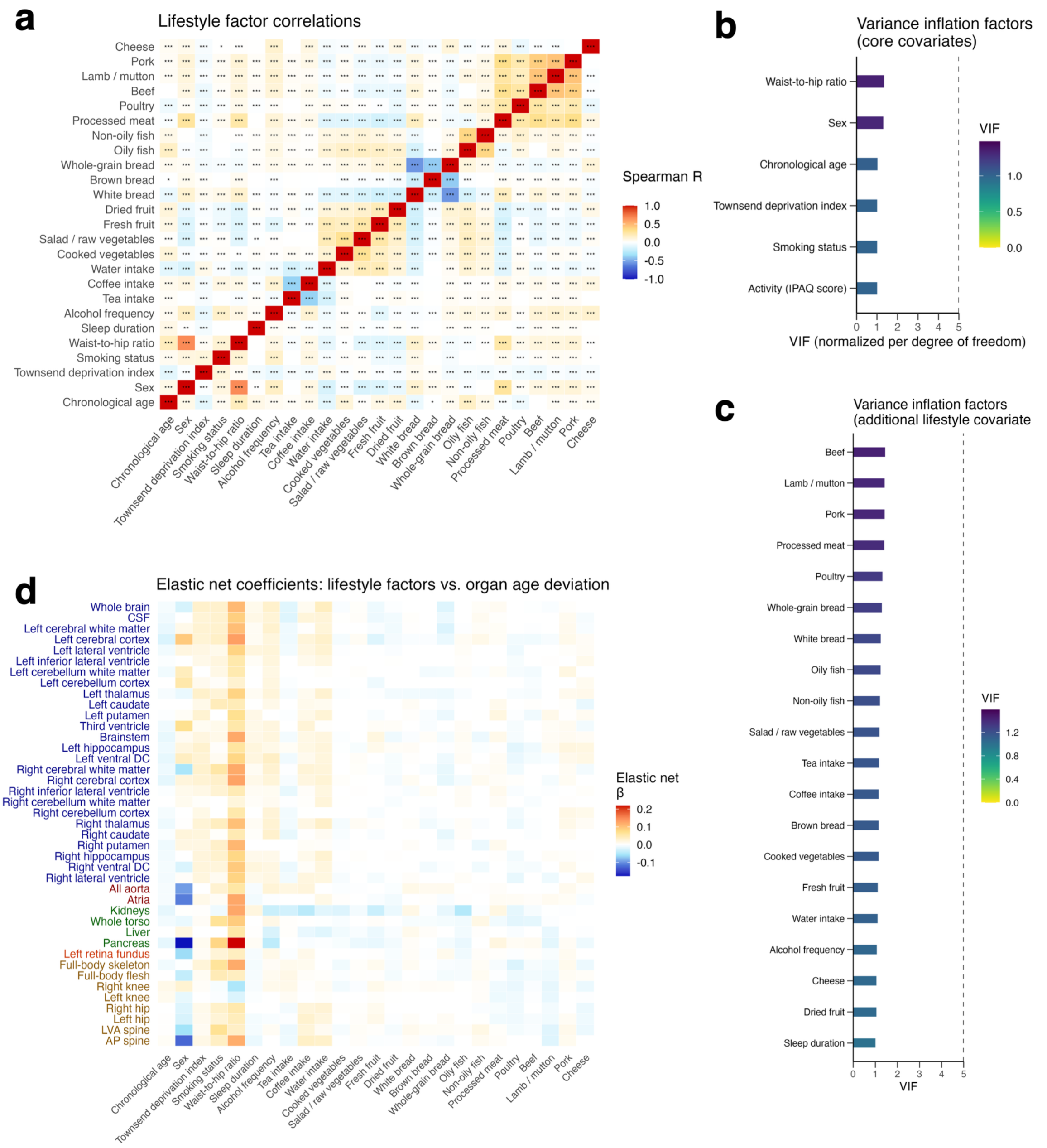


**Figure S7: Additional organ aging lifestyle factor analyses.** Correlation between all lifestyle factors with low (<15%) missingness **(a)**. Collinearity analysis for core confounding covariates **(b)**. Collinearity analysis for all lifestyle factors with low (<15%) missingness **(c)**. Elastic net lifestyle factor analysis heatmap depicting elastic net coefficients for each lifestyle factor and each image model **(d)**. Adjusted p-value < 0.001 = ***, adjusted p-value < 0.01 = **, adjusted p-value < 0.05 = *.
